# A systematic review and meta-analysis of participant characteristics in the prevention of gestational diabetes: a summary of evidence for precision medicine

**DOI:** 10.1101/2023.04.16.23288650

**Authors:** Siew Lim, Wubet Worku Takele, Kimberly K Vesco, Leanne Redman, ADA PMDI GDM prevention working group, Jami Josefson

**Affiliations:** Eastern Health Clinical School, Monash University, Melbourne, Victoria, Australia; Kaiser Permanente Northwest, Kaiser Permanente Center for Health Research, USA; Pennington Biomedical Research Center, Baton Rouge, LA, USA; Northwestern University/ Lurie Children’s Hospital of Chicago, USA; Adelaide Medical School, The University of Adelaide, Australia; Ann & Robert H. Lurie Children’s Hospital of Chicago, Chicago, IL, USA; Global Health Institute, University of Antwerp, 2160 Antwerp, Belgium; Department of Nutrition and Dietetics, Monash University, Melbourne, Victoria, Australia; Madras Diabetes Research Foundation, Deakin University; The University of Adelaide, Adelaide Medical School, Faculty of Health and Medical Sciences; The University of Adelaide; Monash Centre for Health Research and Implementation, Monash University, Clayton, VIC, Australia; Adelaide Medical School, Faculty of Health and Medical Sciences, The University of Adelaide University of Newcastle; School of Agriculture, Food and Wine, University of Adelaide; Department of Women and Children’s health, King’s College London, London, United Kingdom

**Author notes:** **Conflict of interest**. All authors declare no conflict of interest.

## Abstract

**Background and aims:** Precision prevention involves using the unique characteristics of a particular group to determine their responses to preventive interventions. This study aimed to systematically evaluate the participant characteristics associated with interventions in gestational diabetes mellitus (GDM) prevention.

**Methods:** We searched MEDLINE, EMBASE, and Pubmed to identify lifestyle (diet, physical activity, or both), metformin, myoinositol/inositol and probiotics interventions of GDM prevention published up to May 24, 2022.

**Results:** From 10347 studies, 116 studies (n=40940 women) were included. Physical activity resulted in greater GDM reduction in participants with a normal body mass index (BMI) at baseline compared to obese BMI (risk ratio, 95% confidence interval: 0.06 [0.03, 0.14] vs 0.68 [0.26, 1.60]). Diet and physical activity interventions resulted in greater GDM reduction in participants without polycystic ovary syndrome (PCOS) than those with PCOS (0.62 [0.47, 0.82] vs 1.12 [0.78-1.61]) and in those without a history of GDM than those with unspecified history (0.62 [0.47, 0.81] vs 0.85 [0.76, 0.95]). Metformin interventions were more effective in participants with PCOS than those with unspecified status (0.38 [0.19, 0.74] vs 0.59 [0.25, 1.43]), or when commenced preconception than during pregnancy (0.22 [0.11, 0.45] vs 1.15 [0.86-1.55]). Parity, history of having a large-for-gestational-age infant or family history of diabetes had no effect.

**Conclusions:** GDM prevention through metformin or lifestyle differs according to some individual characteristics. Future research should include trials commencing preconception and provide results stratified by participant characteristics including social and environmental factors, clinical traits, and other novel risk factors to predict GDM prevention through interventions.

**Plain language summary:** Precision prevention involves using a group’s unique context to determine their responses to preventive interventions. This study aimed to evaluate the participant characteristics associated with interventions in GDM prevention. We searched medical literature databases to identify lifestyle (diet, physical activity), metformin, myoinositol/inositol and probiotics interventions. A total of 116 studies (n=40903 women) were included. Diet and physical activity interventions resulted in greater GDM reduction in participants without polycystic ovary syndrome (PCOS) and those without a history of GDM. Metformin interventions resulted in greater GDM reduction in participants with PCOS or when started during the preconception period. Future research should include trials starting in the preconception period, and provide results stratified by participant characteristics to predict GDM prevention through interventions.

## INTRODUCTION

Gestational diabetes mellitus (GDM) is characterized by glucose intolerance first identified during pregnancy and is associated with perinatal and long-term adverse health outcomes in both the pregnant individual and the offspring. The physiologic reduction of insulin sensitivity during pregnancy is the hallmark metabolic feature that leads to the onset of glucose intolerance and GDM (1). Established risk factors for GDM include previous GDM, older maternal age, parity, overweight/obese body mass index (BMI), and family history of diabetes (2, 3). GDM increases perinatal complications including preeclampsia, operative deliveries, stillbirth, neonatal risks of large-for-gestational age, hypoglycemia, and respiratory distress syndrome (4). GDM confers an increased lifetime risk of type 2 diabetes mellitus for both mother and offspring (5, 6). GDM rates vary considerably, with geographic differences and varying diagnostic criteria accounting for the 1- 30% incidence (7). Nonetheless, rates of GDM are increasing across all populations (8, 9), commensurate to worldwide increasing rates of overweight and obesity.

Prevention of GDM involves reducing hyperglycemia and insulin resistance, factors that are also highly correlated with obesity (10, 11). Weight reduction prior to pregnancy and prevention of excessive gestational weight gain (GWG) are important features of diabetes prevention (12, 13). Insulin resistance is affected by a number of factors: weight, lifestyle, exercise, diet and supplements. Several meta-analyses of randomized controlled trials (RCTs) investigating lifestyle interventions have reported on diet and physical activity interventions, metformin, and supplements as either primary GDM prevention strategies or secondary prevention strategies for trials targeting weight management and/or reduction as a primary outcome. Results of these meta-analyses have not been unanimous in the reporting of findings suggesting heterogeneity in the intervention response, perhaps due to the characteristics of the study population, and/or the timing and type of intervention (14–17). Variations in the characteristics of the study population such as overweight/obesity, age, and polycystic ovary syndrome (PCOS) could be a potential source of confounding. To date, there has not been a comprehensive meta-analysis of GDM prevention, accounting for participant characteristics to inform precision medicine.

The field of precision medicine recognizes that examining the heterogeneity of individual responses to intervention is important for both optimizing health-enhancing interventions and minimizing exposure to specific risk factors to delay or prevent the onset of a given disease (18, 19).

The *Precision Medicine in Diabetes Initiative* (PMDI) was established in 2018 by the American Diabetes Association (ADA) in partnership with the European Association for the Study of Diabetes (EASD). The ADA/EASD PMDI includes global thought leaders in precision diabetes medicine who are working to address the burgeoning need for better diabetes prevention and care through precision medicine (18). This Systematic Review and subsequent meta-analysis examined the effectiveness of interventions employing lifestyle modification, metformin, or dietary supplements within the preconception, pregnant and postpartum periods for reducing the risk of developing GDM is written on behalf of the ADA/EASD PMDI as part of a comprehensive evidence evaluation in support of the 2^nd^ International Consensus Report on Precision Diabetes Medicine [crossref Tobias et al, Nat Med]. To inform a precision medicine approach to diabetes prevention, the primary focus of this review was to assess the contribution of various participant characteristics to the effectiveness of these interventions for GDM prevention.

## METHODS

This systematic review and meta-analysis was conducted according to the Preferred Reporting Items for Systematic Reviews and Meta-Analyses (PRISMA) Statement (20). The protocol was registered in the PROSPERO International Prospective Register of Systematic Reviews (CRD42022320513).

### Search Strategy

A comprehensive search strategy was developed by a professional medical librarian (AF) in consultation with the authors (SL, LR, KV, JJ). The search strategy included keywords and Medical Subject Headings (MeSH), as shown in Supplementary Table 1. We searched the following databases: Embase (Elsevier), Ovid Medline, and PubMed from inception to May 24, 2022. Results were limited to Human studies and English-language. Endnote (Clarivate) was used to compile records and remove duplicates. Covidence (Veritas Health Innovation, Melbourne, Australia) was then used for title/abstract screening and full text review. Hand-searches including the reference list of related reviews were also examined for additional eligible trials.

### Selection criteria

RCTs and non-randomized control trials (non-RCTs) investigating the effects of lifestyle (diet, exercise, or both), metformin, or dietary supplements (fish oil, myoinositol/inositol, probiotics) on prevention of GDM in women of childbearing age including preconception cohorts were included (Supplementary Table 2). Interventions included lifestyle (diet, exercise, or a combination of these interventions), metformin and dietary supplements. Control conditions include usual care or minimal intervention (no more than a single intervention session in the case of diet and exercise interventions). Studies without a control group (usual care or placebo), those that did not report GDM, observational studies, editorials, commentaries, conference abstracts, reviews, meta-analyses and study protocols were excluded. Titles and abstracts were evaluated independently and in duplicate to identify articles for full-text review. Full-text review was conducted independently and in duplicate with reasons for exclusion recorded. Discrepancies were resolved by consensus by two or more authors.

### Data Extraction

Data were extracted using an extraction template developed for this study with GDM as the primary outcome. Study characteristics (authors, year of publication, country, setting, sample size, design, diagnostic criteria, diagnosis time point, intervention commencement, and outcome of interest), participant characteristics (age, race/ethnicity, BMI, education status, employment status, parity, prior GDM, smoking status and other medical history), intervention type (diet, physical activity, diet and physical activity, metformin, types of dietary supplement), and outcome of intervention (GDM incidence) were extracted. This list of study characteristics was determined based on known GDM risk factors and other relevant factors identified by the precision medicine report (19). Authors were contacted for missing information. One author conducted the data extraction, and a second author conducted a 10% sub-sample data extraction to establish reliability. An agreement of 89% was achieved between the two authors with discussion to resolve discrepancies.

### Quality Assessment

Quality of the included studies was critically appraised using a relevant tool for each study design. The Revised Cochrane Risk of Bias Tool for Randomized Trials (RoB 2.0) (21) was used for RCTs to assess bias arising from the randomization process, deviations from the protocol, missing data, measurement of the outcome and selective reporting. The ROBINS-I tool was used for non-RCTs to assess bias from confounding, participant selection, classification of interventions, missing data, deviations from intended interventions, measurement of outcomes, and selection of reported results (22). Two reviewers independently conducted the methodological quality and bias assessment for individual studies. Differences were resolved by consensus.

The GRADE process (Grading of Recommendations Assessment, Development and Evaluation), which rates the quality of evidence from a study in a systematic approach was conducted for the primary outcome of GDM (23). Risk of bias, along with consistency, directness, precision and publication bias were considered for GRADE appraisal to determine the quality of evidence.

### Statistical Analysis

The outcome was the incidence of GDM. Data were pooled and GDM incidence were expressed as risk ratios (RR) with 95% confidence intervals (CI). Heterogeneity between studies was assessed by the *I*^2^ test where *I*^2^ >50% indicated substantial heterogeneity. Potential sources of heterogeneity by participant characteristics (e.g. obesity, age, PCOS) were explored through subgroup analyses and meta-regression, as conducted in other systematic reviews and meta-analyses (24, 25). Significant (p<0.05) Egger’s test and funnel plot (asymmetry) was used to declare publication bias. Estimates (RR) were pooled using random-effects model with the DerSimonian and Laird estimator (26).

Sensitivity analyses were conducted by excluding non-randomized controlled trials. All analyses were conducted in Stata Version 17 (STATA Corporation, College Station, Texas, USA).

## RESULTS

We screened 10,347 records for eligibility and 434 records were reviewed as full texts (Figure 1). Overall, 130 articles were deemed eligible representing 116 unique studies and were included in the meta-analysis. Reasons for exclusion are as shown in Figure 1, including lack of an appropriate control group where the only difference between the treatment and control group was with or without the interventions of interest, no GDM outcome, active intervention in the control group, and others (Figure 1).

**Figure 1.**
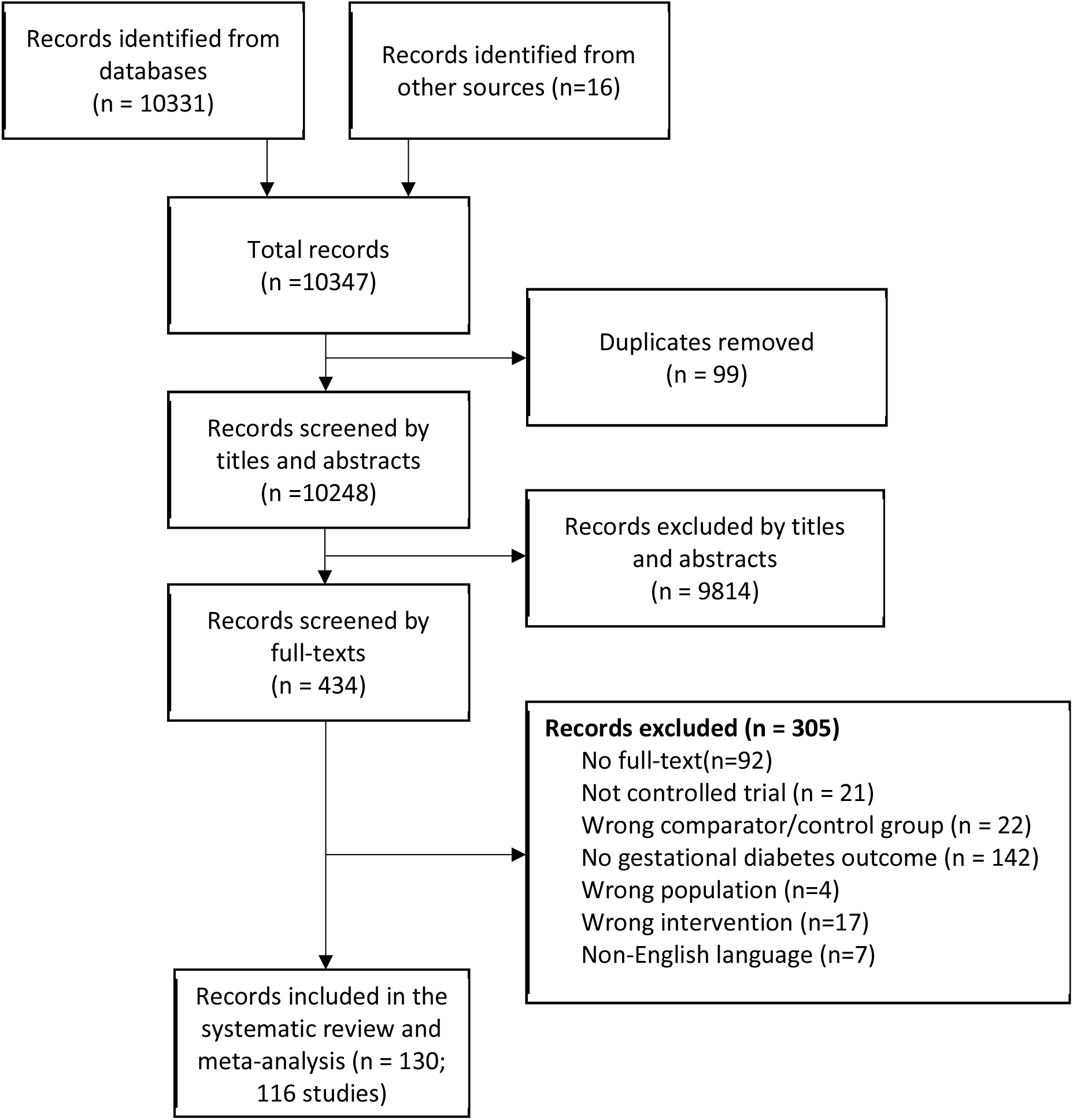
PRISMA flow diagram

### Study characteristics

A summary of the characteristics of included studies are shown in Table 1. Studies were published from 1997 to 2022. Sample sizes ranged from 31 to 4,631. Of the included studies, 92 (79%) involved lifestyle (diet, physical activity, or both), 13 (11%) involved metformin, and 12 (10%) involved dietary supplement interventions. One study included a comparison of lifestyle, probiotics with diet and control (27) and was included as both a lifestyle and dietary supplement study in the meta-analysis. Types of lifestyle intervention included diet only (n = 16), physical activity only (n = 17) or a combination of diet and physical activity (n = 59). The types of dietary supplement interventions included myoinositol/inositol (n = 7), probiotics-only (n = 3), probiotics coupled with diet (n=1), and probiotics with fish oil (n = 1). Interventions commenced from preconception to 26 weeks gestation.

**Table 1.**
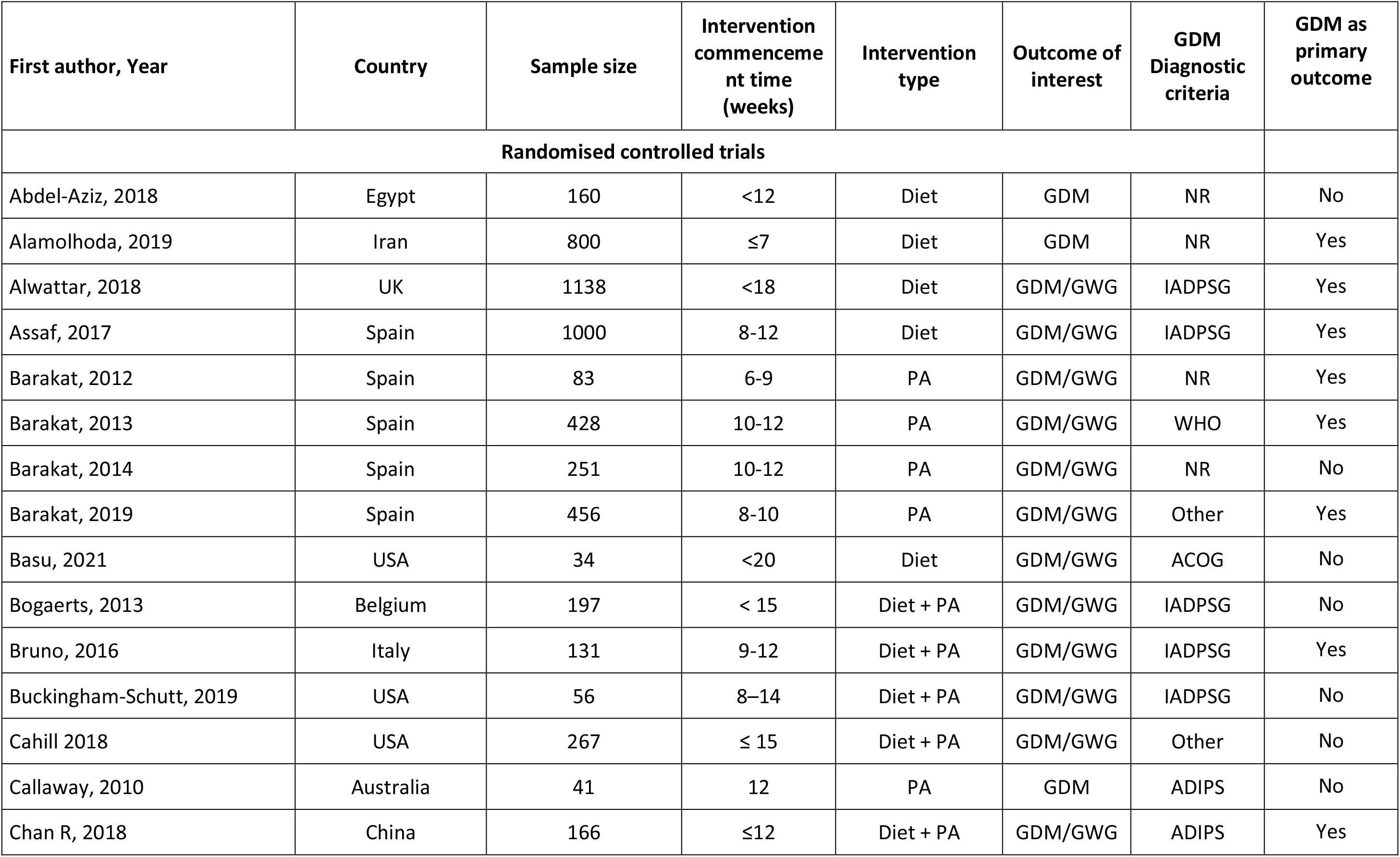
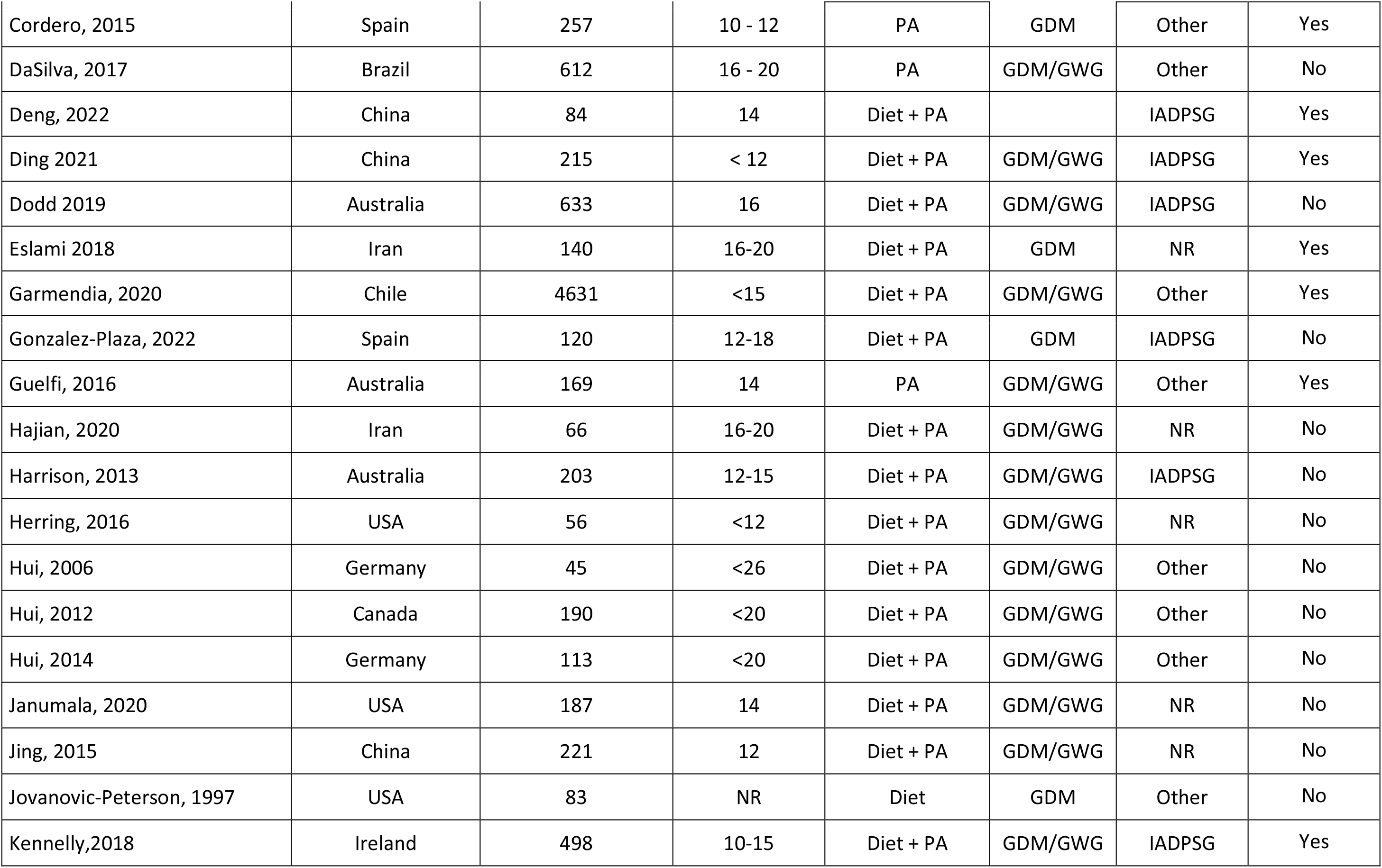
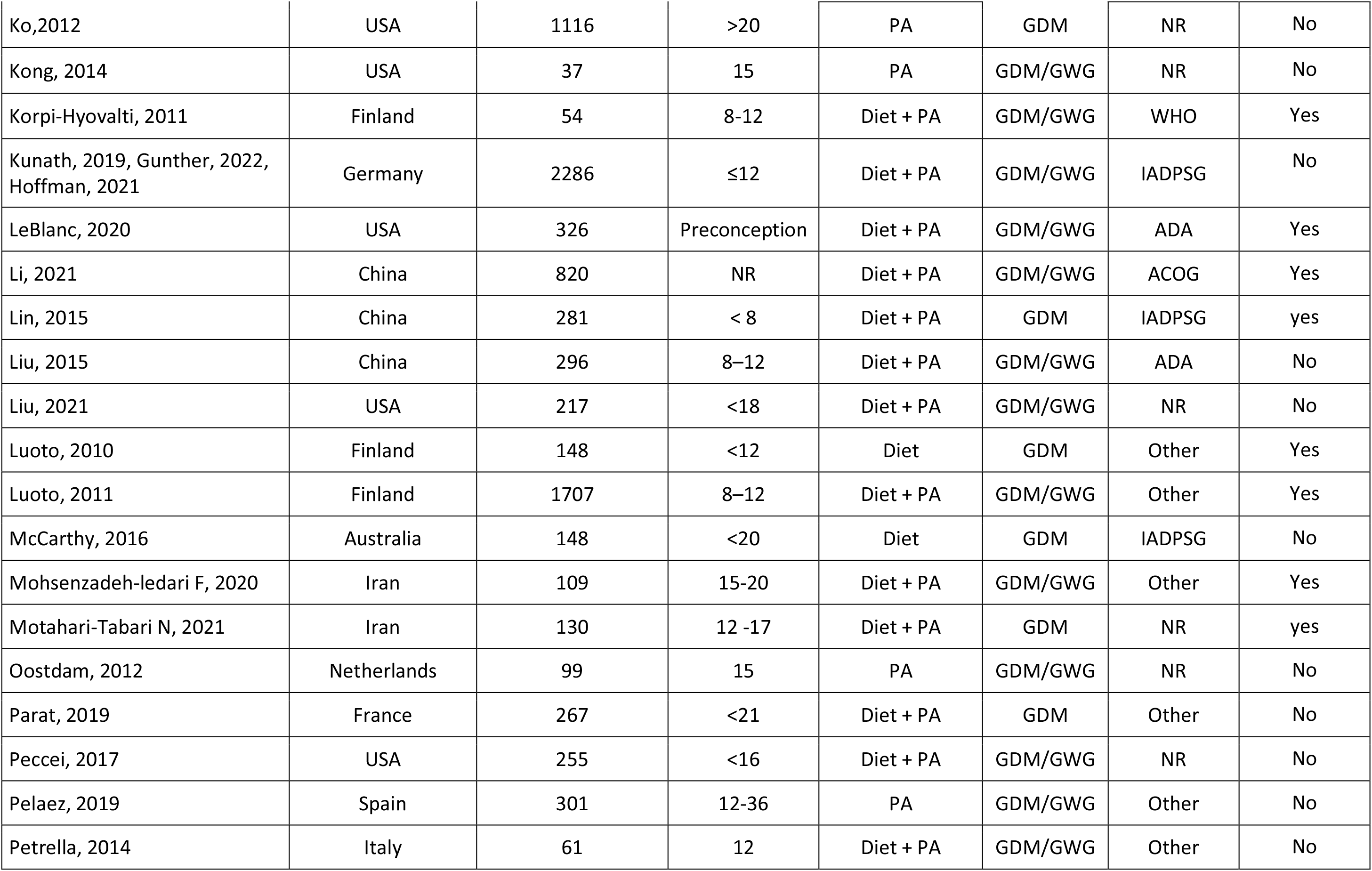
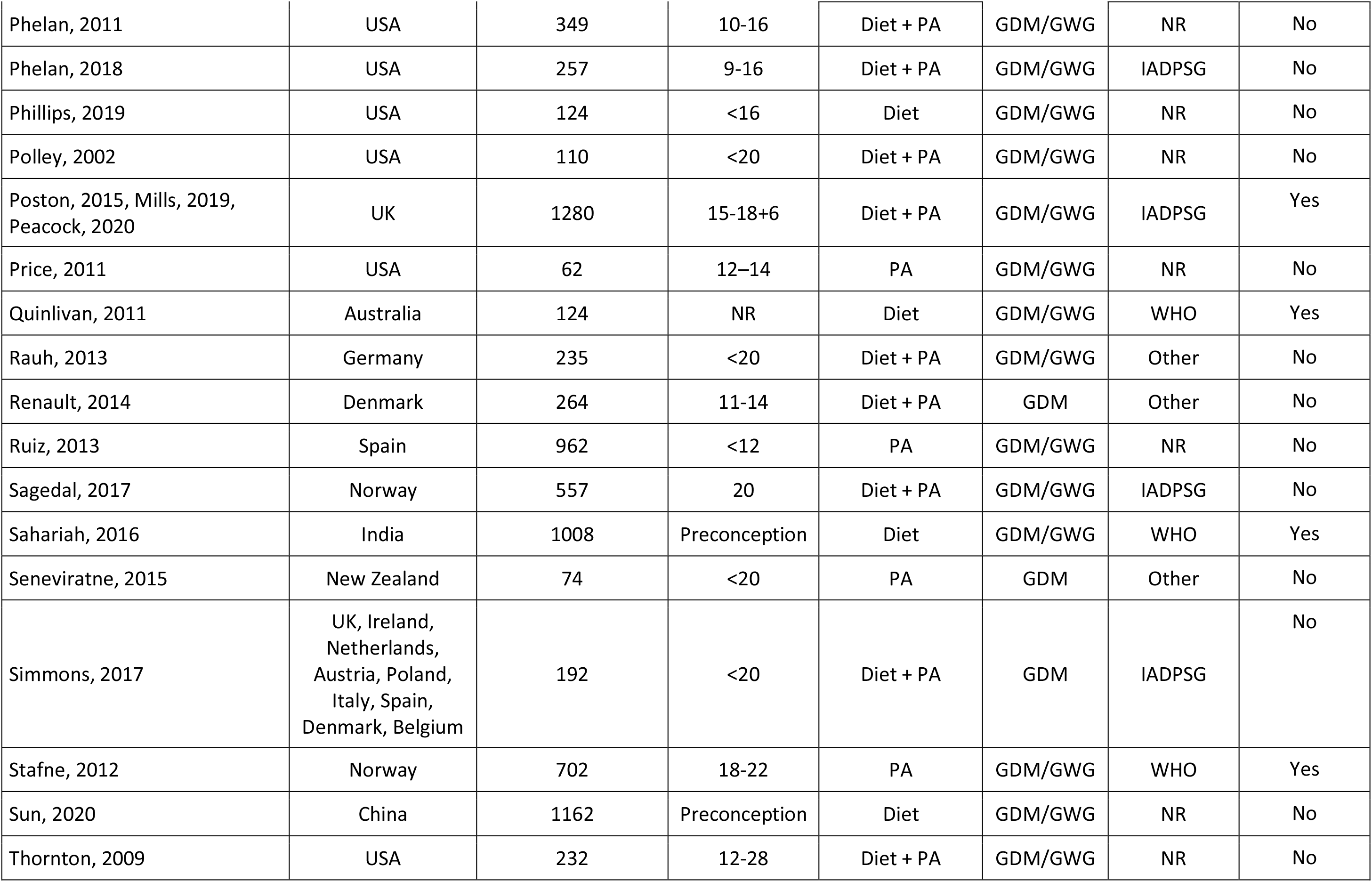
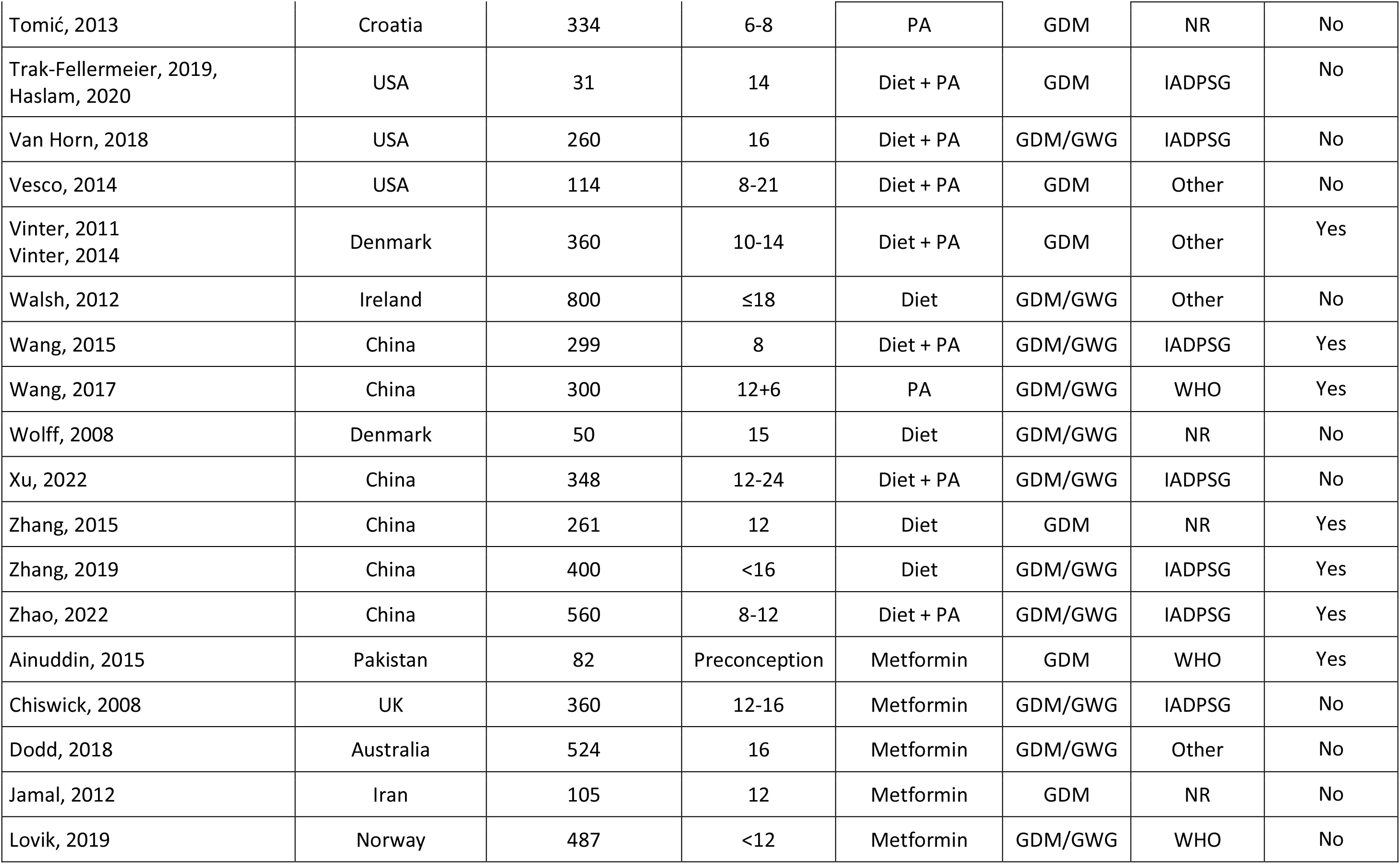
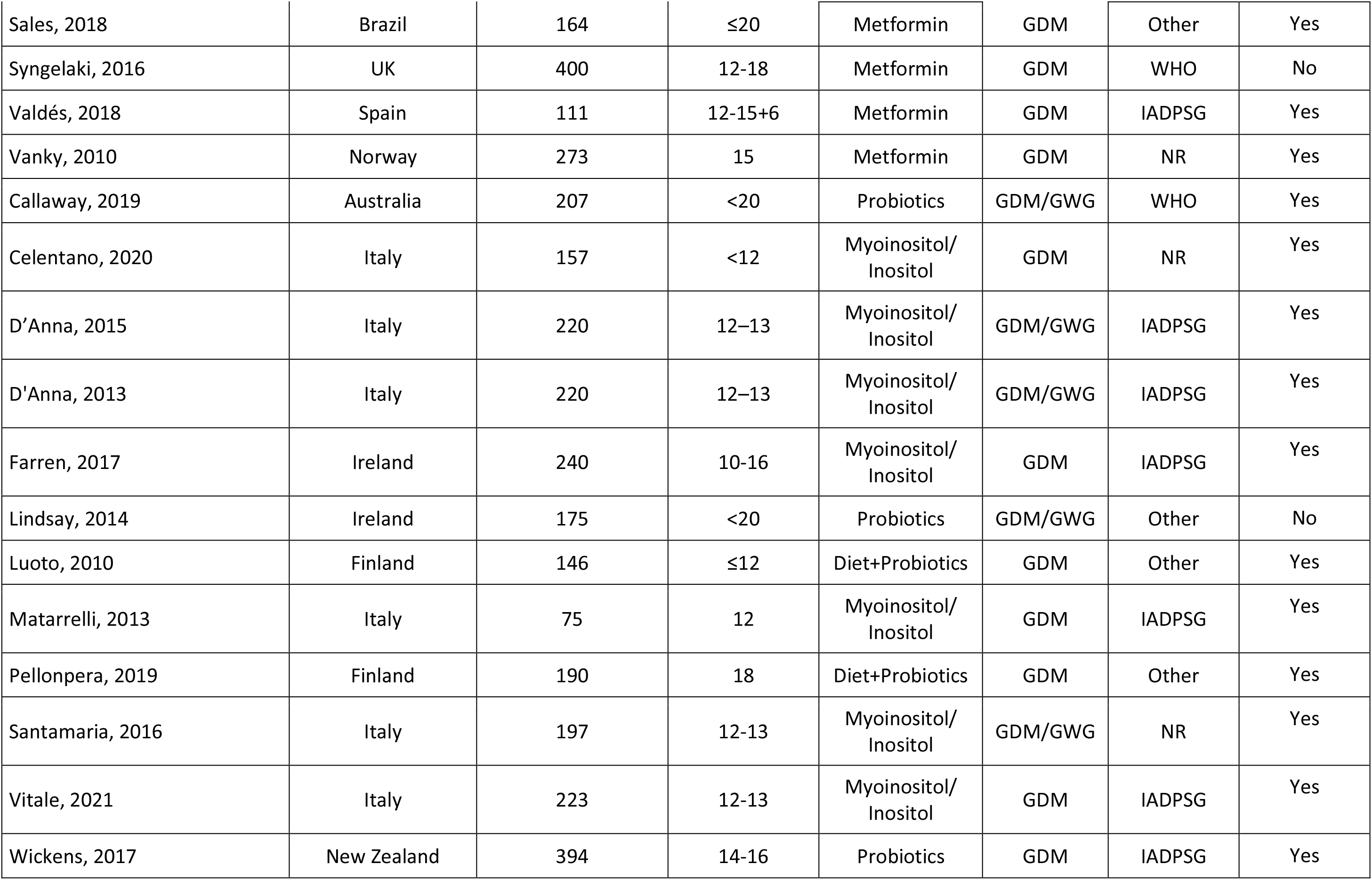
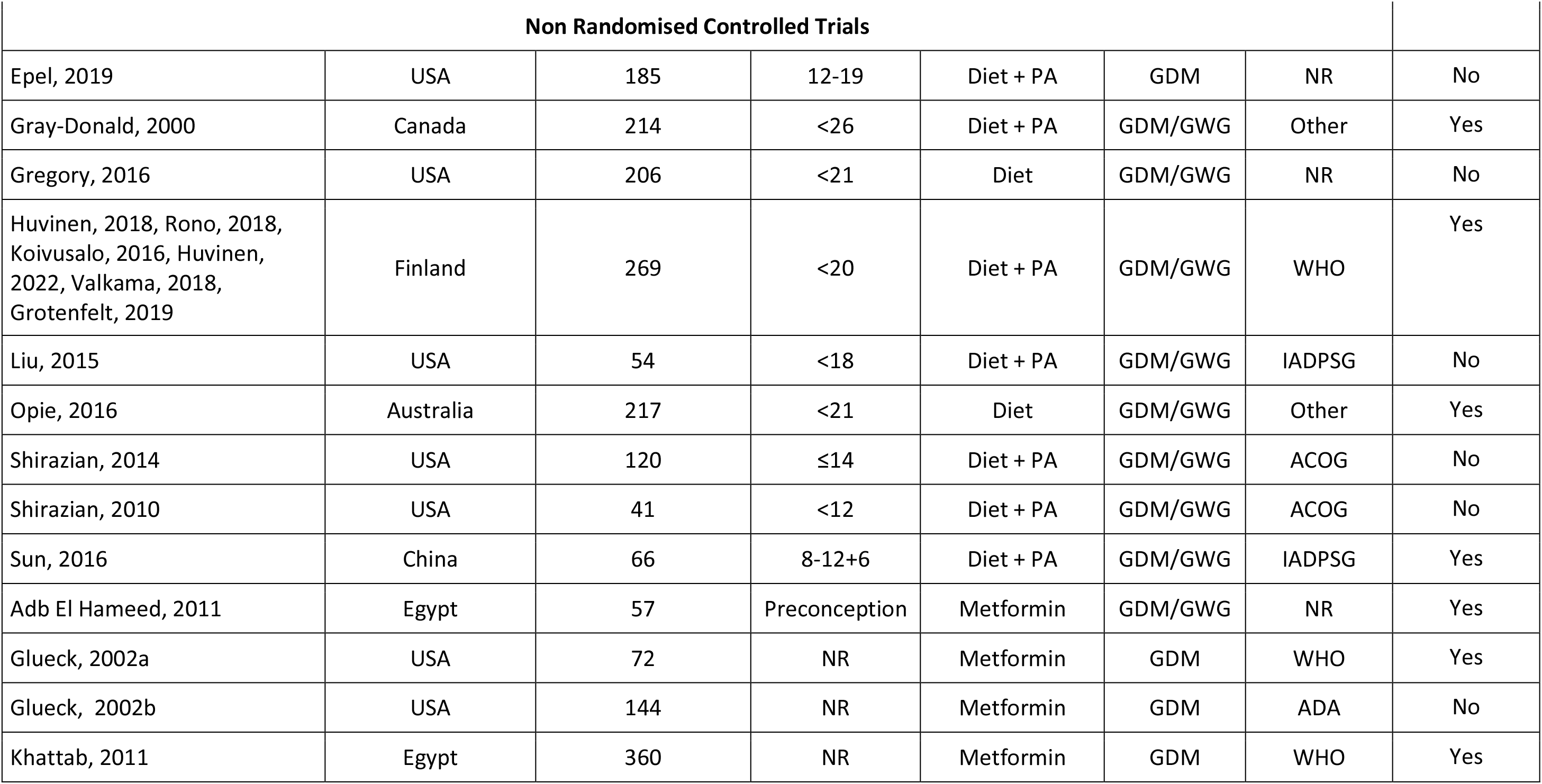
Summary characteristics of included studies

**Table 2.** Meta-analysis of the effect of lifestyle, metformin or dietary supplement on gestational diabetes prevention

| Intervention type | Number of studies | Number of participants | RR (95% CI) | I<sup>2</sup> (%) | Certainty by GRADE | Downgrade explanations |
| --- | --- | --- | --- | --- | --- | --- |
| Diet | 16 | 7303 | 0.75 (0.62-0.90) | 39 | Moderate | Most studies have Some Concerns or High risk of bias |
| Physical activity | 17 | 6243 | 0.69 (0.55-0.85) | 26 | Moderate | Most studies have Some Concerns or High risk of bias |
| Diet and physical activity | 59 | 22026 | 0.82 (0.73-0.91) | 47 | Low | Most studies have Some Concerns or High risk of bias; High levels of heterogeneity |
| Metformin | 13 | 3120 | 0.66 (0.47-0.93) | 73 | Very low | Most studies have Some Concerns or High risk of bias; High levels of heterogeneity; Egger test suggests significant publication bias |
| Myoinositol | 7 | 1313 | 0.39 (0.23-0.66) | 79 | Very low | Most studies have Some Concerns or High risk of bias; Very high levels of heterogeneity |
| Probiotics | 5 | 1104 | 0.88 (0.52-1.47) | 74 | Very low | Most studies have Some Concerns or High risk of bias; Very high levels of heterogeneity; Pooled CI crosses 1.0 suggesting imprecision |

The definition of the participant characteristics is shown in Supplementary Table 3. Detailed description of the participant characteristics of the included studies is as shown in Supplementary Table 4. A total of 105 studies commenced the intervention during pregnancy, six studies reported commencing the intervention prior to pregnancy while five studies did not provide information on pregnancy status at recruitment. Two studies were conducted in women who were nulliparous. A number of studies included only participants with certain medical conditions or medical history: overweight (BMI 25-29.9 kg/m^2^) or obesity (BMI>29.9 kg/m^2^) (n = 57), PCOS (n = 9), prediabetes (n = 1) or family history of diabetes (n = 2); whereas some studies excluded participants with certain medical conditions or medical history: hypertension (n = 27), prediabetes (n = 52), PCOS (n = 3), history of stillbirth (n = 2), family history of diabetes (n = 2), previous macrosomia infant (n = 3), past history of gestational diabetes (n = 20), hypertensive disorders of pregnancy (n = 4), history of cardiovascular disease (n = 14), smoking (n = 11. The mean age of the participants ranged from 25 to 34 years, baseline mean BMI ranged from 21 to 39 kg/m^2^. Of the included studies, 25 (22%) had mostly participants with tertiary education (as defined in Supplementary Table 3and 25 (22%) had mostly participants in employment (Supplementary Table 3). Of the 54 studies which reported race, 18 studies were predominantly conducted among White participants, 10 were predominantly conducted among non-White participants, and 26 among mixed populations.

### Meta-analysis

#### Lifestyle Interventions

Meta-analysis of all lifestyle interventions showed a significant reduction in the risk of GDM with moderate heterogeneity (RR 0.78, 95%CI 0.72, 0.85, *I^2^*=45).

##### Diet-only

In the random effects model, the diet-only interventions showed a significant reduction in the risk of GDM (RR 0.75, 95%CI 0.62, 0.90, 16 studies, *I^2^*=39, moderate-quality evidence) with moderate heterogeneity.

Risk differences in the subgroup and meta-regression analyses by other participant characteristics such as sociodemographic (e.g. educational status) and medical history (e.g. prediabetes and hypertension) was not observed (Table 3 and 4).

**Table 3.**
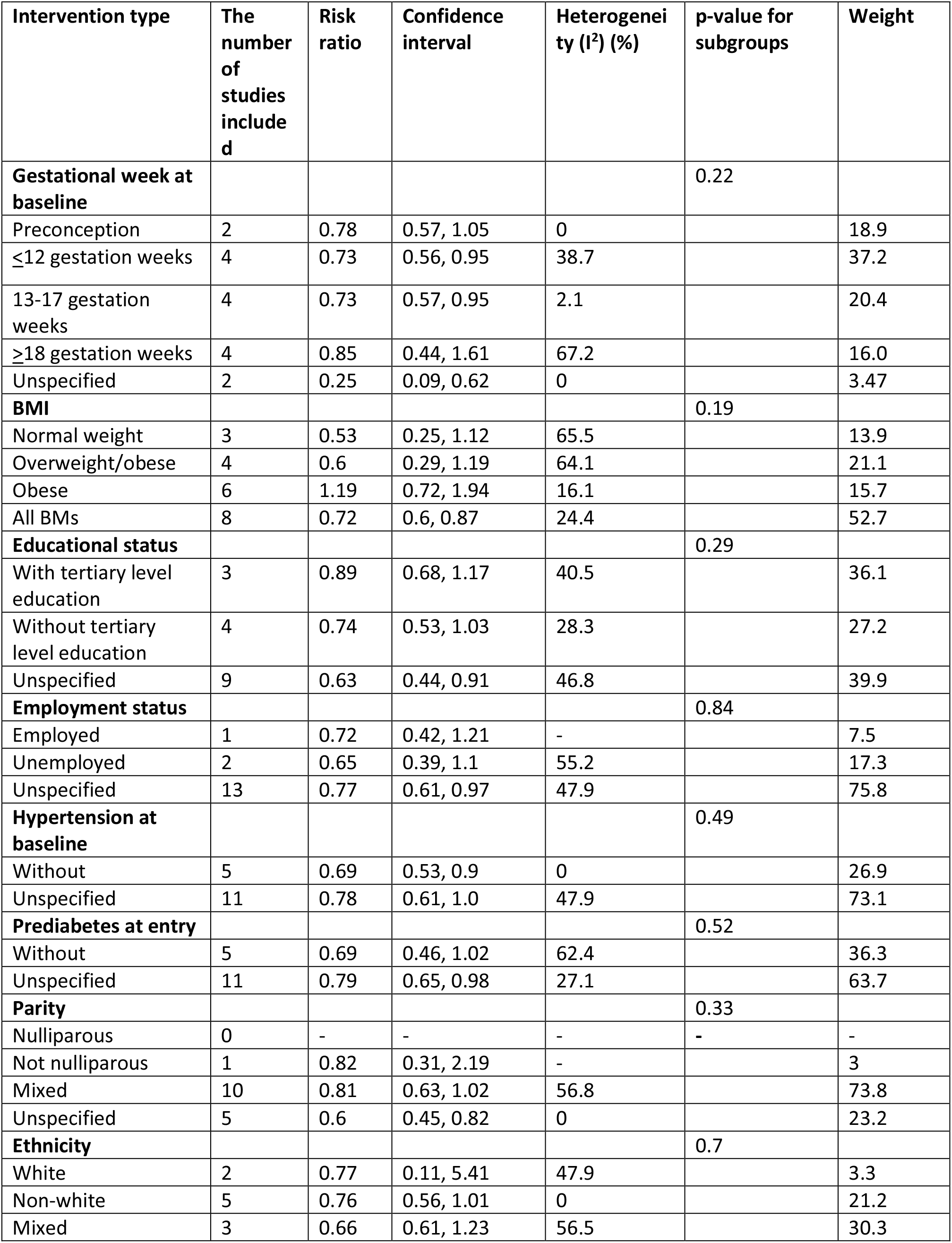
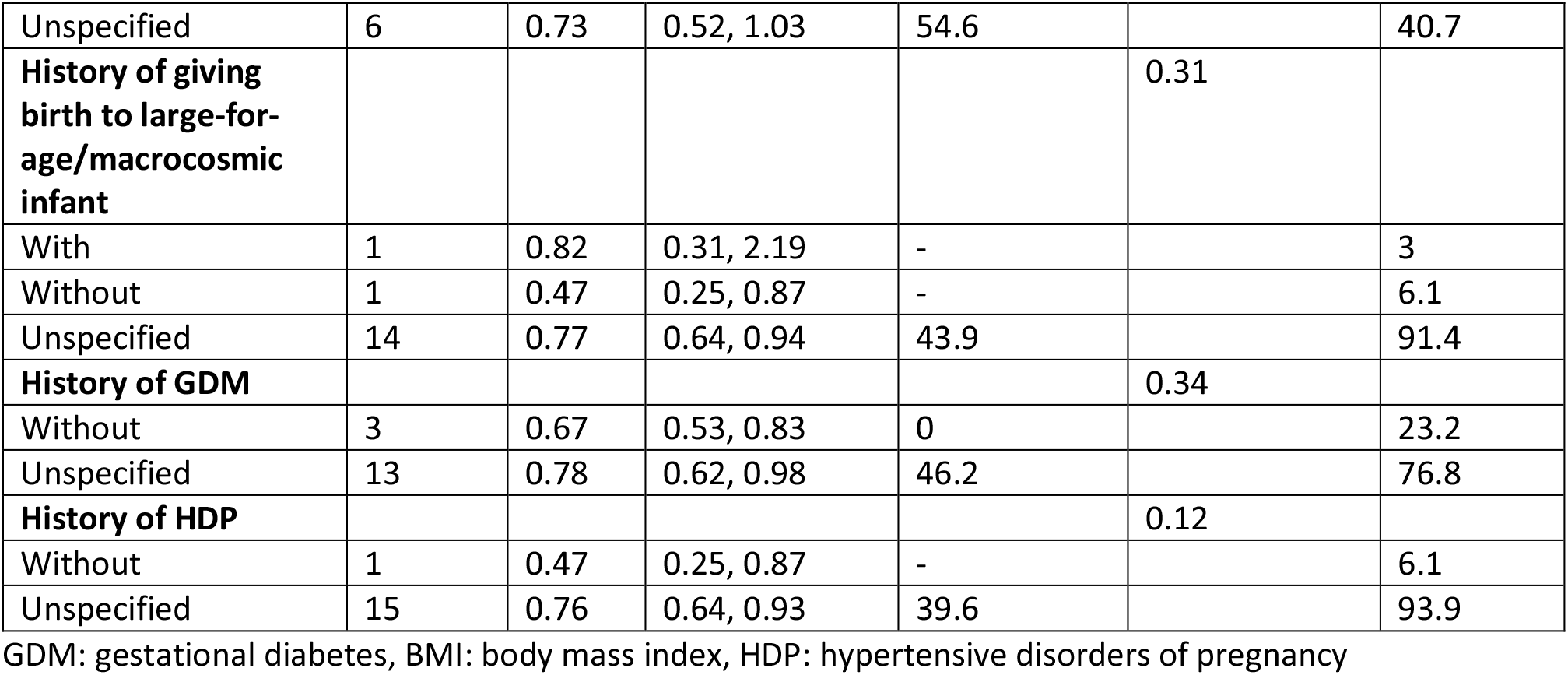
Subgroup analysis of dietary interventions for gestational diabetes prevention, by participant characteristics

##### Physical activity-only

Meta-analysis of all physical activity-only interventions showed a significant reduction in the risk of GDM (RR 0.69, 95%CI 0.55, 0.85, 17 studies, *I^2^*=26, moderate-quality evidence) with moderate heterogeneity.

Subgroup analyses showed that physical activity-only interventions resulted in greater reduction in risk for GDM in studies involving women with normal BMI compared with other BMI groups (Table 5).

**Table 4.** Meta-regression of interventions to prevent gestational diabetes, by participant characteristics

| Intervention and covariate | Coefficient (95% CI) | P-value | Number of studies |
| --- | --- | --- | --- |
| **Dietary intervention** |  |  |  |
| Age (years) | 0.00 (-0.08, 0.08) | 0.99 | 15 |
| BMI (kg/m<sup>2</sup>) | 0.03 (-0.03, 0.09) | 0.25 | 10 |
| Systolic blood pressure (mmHg) | 0.03 (-0.19, 0.25) | 0.67 | 4 |
| Fasting blood glucose (FBG) (mg/dL) | 0.12 (-1.4, 1.64) | 0.51 | 3 |
| **Physical activity intervention** |  |  |  |
| Age (years) | 0.00 (-0.12, 0.12) | 0.99 | 17 |
| BMI (kg/m<sup>2</sup>) | 0.59 (-0.04, 0.16) | 0.22 | 15 |
| Systolic blood pressure (mmHg) | 0.10 (-1.31, 1.33) | 0.93 | 3 |
| HDL(mg/dl) | 0.00 (-0.22, 0.23) | 0.85 | 3 |
| LDL(mg/dl) | 0.00 (0.16, 0.17) | 0.82 | 3 |
| TG (mg/dl) | 0.00 (-0.09, 0.1) | 0.93 | 3 |
| Fasting blood glucose (FBG) (mg/dL) | -0.03 (-0.2, 0.15) | 0.64 | 5 |
| **Combined (diet and physical activity) intervention** |  |  |  |
| Age (years) | -0.50 (-0.10, -0.03) | **0.03** | 55 |
| BMI (kg/m<sup>2</sup>) | -0.01 (-0.03, 0.01) | 0.47 | 50 |
| Systolic blood pressure (mmHg) | -0.01 (-0.06, 0.04) | 0.6 | 9 |
| HDL(mg/dl) | -0.01 (-0.01, 0.03) | 0.23 | 6 |
| LDL(mg/dl) | 0.01 (-0.01, 0.02) | 0.19 | 6 |
| TG (mg/dl) | -0.00 (-0.1, 0.00) | 0.23 | 6 |
| Fasting blood glucose (FBG) (mg/dL) | -0.00 (-0.01, 0.01) | 0.69 | 17 |
| **Metformin intervention** |  |  |  |
| Age(years) | -0.52 (-1.02, -0.02) | **0.04** | 8 |
| BMI (kg/m<sup>2</sup>) | 0.12 (-0.12, 0.35) | 0.26 | 7 |
| Fasting blood glucose (FBG) (mg/dL) | -0.09 (-0.17, -0.03) | **0.02** | 7 |
| **Myoinositol/inositol intervention** |  |  |  |
| Age(years) | 0.11(-0.39, 0.61) | 0.54 | 5 |
| BMI (kg/m<sup>2</sup>) | 0.11(-0.6, 0.83) | 0.69 | 6 |

BMI: body mass index, LDL: low-density lipoprotein, HDL: high-density lipoprotein, TG: triglyceride

**Table 5.** Subgroup analysis of physical activity interventions for gestational diabetes prevention, by participant characteristics

| Intervention type | The number of studies included | Risk ratio | Confidence interval | Heterogeneity ($I^2$) (%) | p-value for subgroups | Weight |
| --- | --- | --- | --- | --- | --- | --- |
| **Gestational week at baseline** |  |  |  |  | 0.06 |  |
| Preconception | 0 | - | - | - | - | - |
| ≤12 gestation weeks | 7 | 0.51 | 0.34, 0.75 | 26.7 |  | 36.9 |
| 13-17 gestation weeks | 6 | 0.72 | 0.53, 0.98 | 17 |  | 40.8 |
| ≥18 gestation weeks | 4 | 0.98 | 0.67, 1.42 | 0 |  | 22.3 |
| **BMI** |  |  |  |  | **0.00** |  |
| Normal weight | 1 | 0.06 | 0.03, 0.14 | - |  | 6.7 |
| Overweight/obese | 5 | 0.59 | 0.42, 0.81 | - |  | 25.4 |
| Obese | 1 | 0.68 | 0.29, 1.6 | 0 |  | 6.5 |
| All BMIs | 11 | 0.72 | 0.53, 0.97 | 36.8 |  | 61.4 |
| **Educational status** |  |  |  |  | **0.02** |  |
| With tertiary level education | 1 | 0.54 | 0.37, 0.79 | - |  | 14.8 |
| Without tertiary level education | 7 | 0.59 | 0.46, 0.77 | 0 |  |  |
| Unspecified | 9 | 0.93 | 0.72, 1.19 | 0 |  |  |
| **Employment status** |  |  |  |  | 0.97 |  |
| Employed | 7 | 0.69 | 0.51, 0.89 | 0 |  | 34.1 |
| Unemployed | 0 | - | - | - | - | - |
| Unspecified | 10 | 0.66 | 0.47, 0.93 | 49.7 |  | 65.9 |
| **Hypertension at baseline** |  |  |  |  | 0.34 |  |
| Without | 7 | 0.61 | 0.48, 0.77 | 0 |  | 42.8 |
| Unspecified | 10 | 0.74 | 0.53, 1.03 | 38.3 |  | 57.3 |
| **Prediabetes at entry** |  |  |  |  | 0.52 |  |
| Without | 7 | 0.63 | 0.46, 0.88 | 44.8 |  | 55.3 |
| Unspecified | 10 | 0.74 | 0.54, 1.01 | 14.1 |  | 44.7 |
| **Parity** |  |  |  |  | 0.34 |  |
| Nulliparous | 1 | 0.4 | 0.08, 2.1 | 0 |  | 1.5 |
| Not nulliparous | 1 | 0.12 | 0.02, 0.91 | 0 |  | 1.01 |
| Mixed | 10 | 0.73 | 0.51, 1.04 | 43.7 |  | 57.2 |
| Unspecified | 6 | 0.67 | 0.52, 0.87 | 0 |  | 40.3 |
| **Ethnicity** |  |  |  |  | 0.07 |  |
| White | 1 | 1.01 | 0.7, 1.46 | - |  | 15.3 |
| Non-white | - | - | - | - | - | - |
| Mixed | 5 | 0.8 | 0.53, 1.19 | 0 |  | 20.6 |
| Unspecified | 11 | 0.59 | 0.44, 0.79 | 30.2 |  | 64.1 |
| **History of GDM** |  |  |  |  | 0.07 |  |
| With | 1 | 1.01 | 0.7, 1.46 | - |  | 15.3 |
| Without | 3 | 0.42 | 0.18, 1.02 | 0 |  | 5.2 |
| Unspecified | 14 | 0.66 | 0.53, 0.82 | 16.7 |  | 79.5 |
| **History of HDP** |  |  |  |  | 0.69 |  |
| Without | 1 | 0.87 | 0.26, 2.91 | - |  | 2.9 |
| Unspecified | 16 | 0.68 | 0.54, 0.85 | 30.2 |  | 97.1 |

GDM: gestational diabetes, BMI: body mass index, HDP: hypertensive disorders of pregnancy

##### Diet and physical activity

Meta-analysis of interventions with both diet and physical components showed a significant reduction in the risk of GDM (RR 0.82, 95%CI 0.73, 0.91, 59 studies, *I^2^*=47, low-quality evidence) with significant heterogeneity.

Subgroup analyses showed that diet and physical activity interventions were not effective in preventing GDM in studies involving women with normal weight but were effective in studies involving women with overweight or obesity (Table 6). Diet and physical activity interventions were also more effective in reducing GDM in studies involving women without PCOS compared with those with PCOS, and in studies involving women without a history of GDM compared with those with unspecified history of GDM (Table 6). Meta-regression showed that diet and physical activity interventions had greater reduction in GDM with increasing age (Table 4).

**Table 6.** Subgroup analysis of combined diet and physical activity interventions for gestational diabetes prevention, by participant characteristics

| Intervention type | The number of studies included | Risk ratio | Confidence interval | Heterogeneity ($I^2$) (%) | p-value for subgroups | Weight |
| --- | --- | --- | --- | --- | --- | --- |
| **Gestational week at baseline** |  |  |  |  | 0.07 |  |
| Preconception | 1 | 0.71 | 0.44, 1.13 | - |  | 2.5 |
| ≤12 gestation weeks | 16 | 0.79 | 0.67, 0.93 | 31.4 |  | 31.3 |
| 13-17 gestation weeks | 22 | 0.87 | 0.72, 1.05 | 48.1 |  | 36.5 |
| ≥18 gestation weeks | 18 | 0.82 | 0.68, 0.99 | 41 |  | 28.7 |
| Trimester of enrolment not specified or across more than one trimester | 1 | 0.18 | 0.06, 0.52 | - |  | 0.85 |
| **BMI** |  |  |  |  | **0.04** |  |
| Normal weight | 2 | 1.66 | 0.72, 3.85 | 0 |  | 1.44 |
| Overweight/obese | 26 | 0.71 | 0.59, 0.87 | 57.3 |  | 43.5 |
| Obese | 12 | 0.94 | 0.8, 1.09 | 0 |  | 16.22 |
| All BMs | 21 | 0.77 | 0.63, 0.95 | 68.7 |  | 38.8 |
| **Educational status** |  |  |  |  | 0.7 |  |
| With tertiary level education | 18 | 0.85 | 0.73, 0.99 | 14.6 |  | 30.9 |
| Without tertiary level education | 21 | 0.75 | 0.6, 0.95 | 63 |  | 32.2 |
| Unspecified | 20 | 0.83 | 0.7, 0.98 | 42.1 |  | 36.9 |
| **Employment status** |  |  |  |  | 0.08 |  |
| Employed | 14 | 0.74 | 0.58, 0.96 | 51.7 |  | 23.2 |
| Unemployed | 4 | 0.5 | 0.29, 0.85 | 37.9 |  | 4.9 |
| Unspecified | 41 | 0.88 | 0.78, 0.98 | 39.2 |  | 71.8 |
| **Hypertension at baseline** |  |  |  |  | 0.7 |  |
| Without | 13 | 0.78 | 0.62, 0.98 | 43.7 |  | 21.5 |
| Unspecified | 46 | 0.83 | 0.73, 0.94 | 48.8 |  | 78.5 |
| **Prediabetes at entry** |  |  |  |  | 0.14 |  |
| Without | 30 | 0.89 | 0.79, 0.99 | 26 |  | 54.02 |
| Unspecified | 29 | 0.75 | 0.62, 0.9 | 59.6 |  | 45.98 |
| **Parity** |  |  |  |  | 0.8 |  |
| Nulliparous | 2 | 1.02 | 0.27, 3.82 | 64.4 |  | 3.2 |
| Mixed | 38 | 0.82 | 0.72, 0.93 | 49.1 |  | 70.5 |
| Unspecified | 19 | 0.77 | 0.63, 0.93 | 27.1 |  | 26.3 |
| **PCOS** |  |  |  |  | **0.03** |  |
| With | 1 | 1.12 | 0.78, 1.61 | - |  | 3.1 |
| Without | 3 | 0.62 | 0.47, 0.82 | 0 |  | 6.8 |
| Unspecified | 55 | 0.83 | 0.74, 0.93 | 46 |  | 91 |
| **Ethnicity** |  |  |  |  | **0.61** |  |
| White | 6 | 0.83 | 0.65, 1.07 | 6.6 |  | 11 |
| Non-white | 5 | 0.83 | 0.58, 1.18 | 0 |  | 6.1 |
| Mixed | 15 | 0.92 | 0.76, 1.01 | 12.5 |  | 20.6 |
| Unspecified | 33 | 0.78 | 0.67, 0.91 | 61.5 |  | 62.3 |
| **History of GDM** |  |  |  |  | **0.03** |  |
| Without | 8 | 0.62 | 0.47, 0.81 | 20.5 |  | 12.1 |
| Unspecified | 51 | 0.85 | 0.76, 0.95 | 44.9 |  | 87.9 |

GDM: gestational diabetes, BMI: body mass index, PCOS: polycystic ovary syndrome

#### Metformin

Meta-analysis of all metformin interventions showed a significant reduction in the risk of GDM (RR 0.66, 95%CI 0.47, 0.93, 13 studies, *I^2^*=73, very low-quality evidence) with significant heterogeneity.

Subgroup analyses showed that metformin interventions were more effective when commenced preconception compared with during pregnancy (Table 7). Metformin interventions were also more effective in reducing GDM in studies involving women with PCOS than those with unspecified status, and less effective in studies involving women with a history of GDM than those with unspecified history of GDM (Table 7). Meta-regression showed that metformin interventions were more effective in reducing GDM with increasing age or fasting blood glucose (FBG) (Table 4).

**Table 7.**
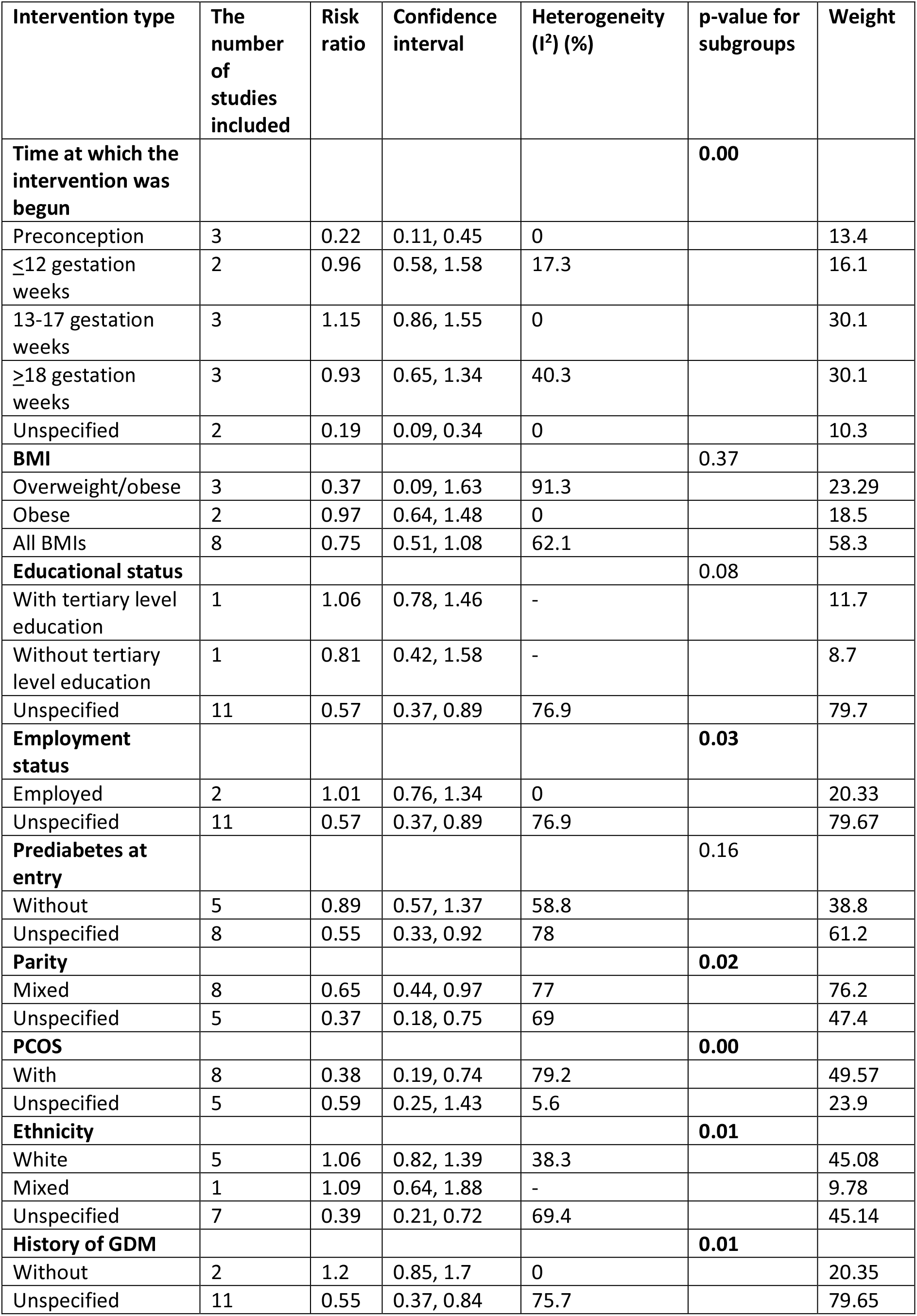
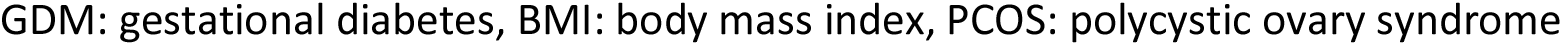
Subgroup analysis of metformin interventions for gestational diabetes prevention, by participant characteristics

| Intervention type | The number of studies included | Risk ratio | Confidence interval | Heterogeneity (I<sup>2</sup>) (%) | p-value for subgroups | Weight |
| --- | --- | --- | --- | --- | --- | --- |
| **Time at which the intervention was begun** |  |  |  |  | **0.00** |  |
| Preconception | 3 | 0.22 | 0.11, 0.45 | 0 |  | 13.4 |
| ≤12 gestation weeks | 2 | 0.96 | 0.58, 1.58 | 17.3 |  | 16.1 |
| 13-17 gestation weeks | 3 | 1.15 | 0.86, 1.55 | 0 |  | 30.1 |
| ≥18 gestation weeks | 3 | 0.93 | 0.65, 1.34 | 40.3 |  | 30.1 |
| Unspecified | 2 | 0.19 | 0.09, 0.34 | 0 |  | 10.3 |
| **BMI** |  |  |  |  | **0.37** |  |
| Overweight/obese | 3 | 0.37 | 0.09, 1.63 | 91.3 |  | 23.29 |
| Obese | 2 | 0.97 | 0.64, 1.48 | 0 |  | 18.5 |
| All BMIs | 8 | 0.75 | 0.51, 1.08 | 62.1 |  | 58.3 |
| **Educational status** |  |  |  |  | **0.08** |  |
| With tertiary level education | 1 | 1.06 | 0.78, 1.46 | - |  | 11.7 |
| Without tertiary level education | 1 | 0.81 | 0.42, 1.58 | - |  | 8.7 |
| Unspecified | 11 | 0.57 | 0.37, 0.89 | 76.9 |  | 79.7 |
| **Employment status** |  |  |  |  | **0.03** |  |
| Employed | 2 | 1.01 | 0.76, 1.34 | 0 |  | 20.33 |
| Unspecified | 11 | 0.57 | 0.37, 0.89 | 76.9 |  | 79.67 |
| **Prediabetes at entry** |  |  |  |  | **0.16** |  |
| Without | 5 | 0.89 | 0.57, 1.37 | 58.8 |  | 38.8 |
| Unspecified | 8 | 0.55 | 0.33, 0.92 | 78 |  | 61.2 |
| **Parity** |  |  |  |  | **0.02** |  |
| Mixed | 8 | 0.65 | 0.44, 0.97 | 77 |  | 76.2 |
| Unspecified | 5 | 0.37 | 0.18, 0.75 | 69 |  | 47.4 |
| **PCOS** |  |  |  |  | **0.00** |  |
| With | 8 | 0.38 | 0.19, 0.74 | 79.2 |  | 49.57 |
| Unspecified | 5 | 0.59 | 0.25, 1.43 | 5.6 |  | 23.9 |
| **Ethnicity** |  |  |  |  | **0.01** |  |
| White | 5 | 1.06 | 0.82, 1.39 | 38.3 |  | 45.08 |
| Mixed | 1 | 1.09 | 0.64, 1.88 | - |  | 9.78 |
| Unspecified | 7 | 0.39 | 0.21, 0.72 | 69.4 |  | 45.14 |
| **History of GDM** |  |  |  |  | **0.01** |  |
| Without | 2 | 1.2 | 0.85, 1.7 | 0 |  | 20.35 |
| Unspecified | 11 | 0.55 | 0.37, 0.84 | 75.7 |  | 79.65 |

GDM: gestational diabetes, BMI: body mass index, PCOS: polycystic ovary syndrome

#### Dietary supplements

##### Myoinositol/Inositol

Meta-analysis of all myoinositol/inositol interventions showed a significant reduction in the risk of GDM (RR 0.39, 95%CI 0.23, 0.66, 7 studies, *I^2^*=79, very low-quality evidence) with significant heterogeneity.

Subgroup analyses showed that myoinositol/inositol interventions were more effective in reducing GDM in studies involving mostly White women compared with women from various ethnic backgrounds (Table 8).

**Table 8.** Subgroup analysis of myoinositol/inositol for gestational diabetes prevention, by participant characteristics

| Intervention type | The number of studies included | RR | Confidence interval | Heterogeneity (I) (%) | p-value for subgroups | Weight |
| --- | --- | --- | --- | --- | --- | --- |
| **Gestational week at which the intervention was begun** |  |  |  |  | 0.87 |  |
| Preconception | 1 | 0.4 | 0.27, 0.59 |  |  | 17.42 |
| ≤12 gestation weeks | 6 | 0.38 | 0.19, 0.74 |  |  | 82.6 |
| 13-17 gestation weeks | 0 | - | - | - | - | - |
| **BMI** |  |  |  |  | 0.88 |  |
| Normal weight | 1 | 0.42 | 0.24, 0.72 | - |  | 16.03 |
| Overweight/obese | 2 | 0.3 | 0.15, 0.59 | 54.2 |  | 29.6 |
| All BMIs | 2 | 0.35 | 0.02, 5.15 | 92.8 |  | 24.7 |
| Obese | 2 | 0.4 | 0.28, 0.58 | 0 |  | 29.6 |
| **Hypertension** |  |  |  |  | 0.82 |  |
| Without | 1 | 0.42 | 0.24, 0.72 | - |  | 83.9 |
| Unspecified | 6 | 0.38 | 0.2, 0.72 | 82.3 |  | 83.9 |
| **Prediabetes at baseline** |  |  |  |  | 0.07 |  |
| With | 1 | 0.08 | 0.02, 0.31 | - |  | 8.28 |
| Without | 5 | 0.46 | 0.24, 0.88 | 80.9 |  | 74.3 |
| Unspecified | 1 | 0.4 | 0.27, 0.59 | - |  | 17.2 |
| **Ethnicity** |  |  |  |  | 0.00 |  |
| White | 2 | 0.41 | 0.25, 0.69 | 0 |  | 27.2 |
| Mixed | 1 | 1.27 | 0.77, 2.09 | - |  | 16.5 |
| Unspecified | 4 | 0.29 | 0.18, 0.48 | 60.2 |  | 56.4 |
| **History of GDM** |  |  |  |  | 0.81 |  |
| Without | 4 | 0.35 | 0.25, 0.49 | 1.5 |  | 57.85 |
| Unspecified | 3 | 0.41 | 0.13, 1.28 | 90.4 |  | 42.2 |

GDM: gestational diabetes, BMI: body mass index

##### Probiotics

Meta-analysis of all probiotics interventions showed no reduction in the risk of GDM (RR 0.88, 95%CI 0.52, 1.47, 5 studies, *I^2^*=74, very low-quality evidence) with significant heterogeneity. Of these, one study with diet plus probiotics resulted in significant reduction in GDM (RR 0.36, 95%CI 0.18,0.72) (27), however three studies with probiotics-only (28–31) and one study with fish oil-plus probiotics (31) did not reduce GDM (RR ranging from 0.59 to 1.5).

Subgroup analyses and meta-regression were not conducted in probiotics interventions due to small numbers in each type of intervention.

##### Fish oil

The only fish oil study found that it did not reduce GDM (RR 1.09, 95%CI 0.64, 1.85) (31).

##### Sensitivity analysis

After excluding two non-RCT studies, diet-only interventions remained significant in preventing GDM (RR 0.75; 95% CI; 0.64, 0.88; *I^2^*=23). After excluding six non-RCT studies, combined diet and physical activity interventions remained significant in reducing GDM (RR 0.83; 95% CI; 0.74, 0.93; *I^2^*=64.8%). After excluding five non-RCT studies, the effect of metformin on reducing the incidence of GDM was no longer significant (RR 1.05, 95% CI 0.89, 1.23; *I^2^*=45.1).

Sensitivity analysis was not conducted on physical activity-only, probiotics and myoinositol/inositol studies as all were RCTs.

##### Assessment of bias and quality of evidence

Some concerns or high risk of bias were found in 12 (80%) of diet-only, 16 (94%) physical activity-only, 46 (78%) of diet and physical activity, 10 (77%) of metformin, 5 (71%) in myoinositol/inositol, 3 (60%) in probiotics (Supplementary Table 5). Most studies assigned a high risk of bias had insufficient or non-blinding of participants as the main reason.

The quality of evidence rated by the GRADE approach found that the overall quality for diet-only or physical activity-only interventions were moderate, downgraded mainly due to most studies contributing to the outcome having high or some concerns in risk of bias. (Supplementary Table 6). The quality of evidence for diet and physical activity interventions were low, due to risk of bias and inconsistency. The quality of evidence for metformin interventions was very low, due to risk of bias, inconsistency and publication bias. The quality of evidence for myoinositol/inositol interventions was very low, due to risk of bias and inconsistency. The quality of evidence for probiotics interventions was very low due to risk of bias, inconsistency and imprecision.

##### Publication bias

Funnel plot and Egger’s test suggested the presence of small studies publication bias for metformin studies (P=0.001) (Supplementary Figure 1).

No significant publication bias was detected for physical activity-only, diet-only studies, diet and physical activity studies, probiotics, on GDM prevention. (Supplementary Figure 1).

## DISCUSSION

This study aimed to determine the effect of participant characteristics in interventions for GDM prevention. Our analyses showed that lifestyle interventions, metformin or myoinositol/inositol reduced the risk of GDM. For physical activity-only interventions, greater risk reduction for GDM was seen in studies involving women with normal BMI. Combined diet and physical activity interventions were more effective in GDM reduction in those with overweight or obesity, without PCOS, without history of GDM and with increasing age. Metformin interventions were more effective in GDM reduction in women with a history of PCOS and with increasing age and fasting blood glucose. Metformin interventions were also more effective when commenced preconception.

Diet and physical activity interventions were more effective for lowering risk for GDM among women without a history of GDM. This may be due to women who had prior GDM has a history of impaired beta-cell compensatory response during pregnancy followed by possibly continual worsening of insulin sensitivity post-GDM pregnancy (11). Lifestyle modification alone may have limited ability to overcome these impairments in glycemic control in these women. In addition, lower adherence to a healthy diet as lower dietary quality which has been observed among women with a history of GDM compared with women without a history of GDM may have also contributed to this (32). Similarly, we also found that lifestyle interventions more effective in lowering GDM risks in women without a history of PCOS, a population characterised by high insulin resistance (33). In addition to the physiological challenges of insulin resistance, women with PCOS may also face further challenges with adhering to a healthy lifestyle which range from physiological barriers such as alteration in gut hormone regulation to psychological barriers such as a high prevalence of disordered eating in this population (34). Further research is needed to determine if more intensive lifestyle intervention or additional co-intervention such as metformin or supplementation is needed to prevent GDM in women with conditions of high insulin resistance such as a history of prior GDM or PCOS.

Systematic reviews to date have been inconsistent on the effect of metformin on GDM prevention (16, 35), which could be due to heterogeneity in participant characteristics which undermines the power to detect a significant effect within a small number of studies (16, 35). In support of this, past studies with a select population thereby reducing participant heterogeneity, such as studies in women with PCOS, has seen a consistent benefit in GDM prevention with metformin (36, 37). By increasing the number of included studies from 3 in the previous meta-analysis (16) to the current meta-analyses of 13 metformin trials, the current review revealed a significant reduction in GDM with metformin. To further explore the sources of heterogeneity by participant characteristics, our meta-regression additionally found that metformin is more effective in studies involving women with an older mean age at baseline, or in women with higher baseline fasting blood glucose. An increase in age is associated with greater insulin resistance while higher baseline fasting glucose indicates early signs of failure of beta-cell compensatory response in insulin production against chronic insulin resistance (38). The greater benefit of metformin in women with increased insulin resistance is in line with the known mechanisms of metformin, which is to reduce glucose production in the liver, improve peripheral glucose uptake and increase insulin sensitivity (39). Further studies should be conducted to confirm if metformin should be prescribed to prevent GDM in populations at high risk of insulin resistance, including in women of higher age, higher fasting blood glucose, history of GDM and with PCOS, along with healthy antenatal lifestyle advice.

Similar to a previous meta-analysis (15), we found that lifestyle or metformin interventions were more effective in lowering the risk for GDM when commenced preconception or in the first trimester of pregnancy. Earlier initiation of interventions results in a greater duration of intervention exposure prior to GDM diagnosis, while preconception commencement additionally optimises insulin sensitivity prior to the onset of pregnancy-induced insulin resistance. A preconception intervention is in line with the concept of GDM as a chronic metabolic disease. Whilst first identified in pregnancy, women who developed GDM were already on a trajectory of increased cardiometabolic risk prior to pregnancy (40). Future research should focus on providing preconception GDM prevention in those at risk of developing GDM. This is in support of the growing research area of risk prediction of GDM in preconception populations, which will identify the populations which will optimally benefit from preconception GDM prevention (41).

The strength of this review includes the first of its kind to investigate the differential effects of GDM prevention in population groups across a comprehensive range of participant characteristics on all interventions for GDM prevention to identify populations that could optimally benefit from each intervention type. Although most studies were conducted in mixed populations and did not report outcomes according to these subgroups bearing these characteristics, we coded the participant subgroups according to the inclusion and exclusion criteria (e.g. if the study included only women with obesity) to allow for group comparisons. This review also has several limitations. Participant characteristics that seldom feature in the inclusion or exclusion criteria of the included studies, such as parity, yielded a low number of subgroups available for comparisons, despite the known confounding relationship between these characteristics and GDM risks. Significant heterogeneity remained in some subgroups, suggesting that other confounding factors may have contributed to the effect sizes. One of the confounding effects that are difficult to quantify is the changing GDM diagnostic criteria over the years which may have also contributed to the heterogeneity of the participants in the analyses. The findings on lifestyle or metformin on GDM incidence may also be limited by possible publication bias. The certainty of evidence was very low for metformin, myoinositol/inositol and probiotics, and should be interpreted with caution.

## Conclusion

Results from this meta-analysis found that lifestyle, metformin and myoinositol/inositol reduce the risk of GDM. Lower GDM risks were seen when the intervention commenced preconception or in early first trimester. Diet and physical activity interventions may be associated with greater reduction in GDM risks in women with older age or without a history of GDM or PCOS, while metformin may be more effective in preventing GDM in women with older age, having higher fasting blood glucose or with PCOS. Given the greater effectiveness of these interventions in certain groups of individuals, future research on tailored recommendations in precision GDM prevention, replacing the current ‘one-size-fits-all’ approach, is needed. To advance knowledge in precision prevention, future research should include trials commencing preconception and provide results stratified by participant characteristics including social and environmental factors, clinical traits, and other novel risk factors to predict GDM prevention through interventions

## Authors contribution

SL, JJ, LR, KV conceptualised the research question. AF contributed to the search of the articles. SL, MC, JG, NH, LR, JJ, KV, WT, KL, GGU, SC screened the articles, JG, NH, WT, GGU, GL, SJZ, RT, MP, KL,

MB, AQ, HW extracted the data and appraised the studies, WT and SL coded the participant characteristics, WT, GGU and MC contributed to data analysis, all authors contributed to the manuscript.

## Data Availability

All data produced in the present study are available upon reasonable request to the authors

## Acknowledgement

SL is funded by the Australian National Health and Medical Research Council (NHMRC) Fellowship. JW is funded by NHMRC Ideas Grant. WT, MC and GU were funded by the Australian Government Research Training Program Scholarship. LR is funded by the National Institute of Health (5R01DK124806).

## List of Tables and Figures

Supplementary Table 1: Search strategies

Supplementary Table 2: Inclusion criteria of included studies

Supplementary Table 3: Definition of participant characteristics

Supplementary Table 4: Participant characteristics of included studies

Supplementary Table 5: Risk of bias summary

Supplementary Table 6: GRADE summary

Supplementary Figure 1. Funnel plots for lifestyle, metformin and dietary supplement interventions on GDM incidence

## Notes

### Competing Interest Statement

The authors have declared no competing interest.

