## Supplementary for "A systematic review and meta-analysis of participant characteristics in the prevention of gestational diabetes: a summary of evidence for precision medicine"

List of Tables and Figures

Table 1. Summary characteristics of included studies

Table 2. Meta-analysis of the effect of lifestyle, metformin or dietary supplement on gestational diabetes prevention

Table 3. Subgroup analysis of dietary interventions for gestational diabetes prevention, by participant characteristics

Table 4. Meta-regression for gestational diabetes prevention, by participant characteristics

Table 5. Subgroup analysis of physical activity interventions for gestational diabetes prevention, by participant characteristics

Table 6. Subgroup analysis of combined diet and physical activity interventions for gestational diabetes prevention, by participant characteristics

Table 7. Subgroup analysis of metformin interventions for gestational diabetes prevention, by participant characteristics

Table 8. Subgroup analysis of myoinositol/inositol for gestational diabetes prevention, by participant characteristics

Table 9. Subgroup analysis of probiotics for gestational diabetes prevention, by participant characteristics

Figure 1. PRISMA flow diagram

Supplementary Table 1: Search strategies

Supplementary Table 2: Inclusion criteria of included studies

Supplementary Table 3: Definition of participant characteristics

Supplementary Table 4: Participant characteristics of included studies

Supplementary Table 5: Risk of bias summary

Supplementary Table 6: GRADE summary

Supplementary Figure 1. Funnel plots for lifestyle, metformin and dietary supplement interventions on GDM incidence

Supplementary Table 1. Search strategies

| Search Queries |
|---|
| **Medline (Ovid)**<br><br>1. Pregnancy/ or Gravidity/ or Preconception Care/<br>2. (antenatal or (ante* adj2 natal) or childbearing or (child adj2 bearing) or gestation* or family planning services or gravid* or interconcept* or intergestation* or internatal or matern* or periconcept* or preconcept* or pregestation* or prenatal* or prepregn* or ((inter or pre or peri) adj2 (concept* or gestation* or natal or pregnan*))).ti,ab,kw.<br>3. 1 or 2<br>4. Behavior Therapy/ or Cognitive Behavioral Therapy/ or Diet/ or Exercise/ or Health Behavior/ or Health Education/ or Health Promotion/ or Life Style/ or Weight Loss/<br>5. ((behav* adj2 (cognit* or manag* or modif* or therap*)) or CBT or diet* or (health* adj2 (behav* or eat* or educat* or food* or promotion)) or lifestyle or (life adj2 style) or nutrition or physical activit* or (weight adj2 (loss or manag* or reduction or retention))).ti,ab,kw.<br>6. 4 or 5<br>7. Biguanides/ or Metformin/ or Buformin/ or Chlorhexidine/ or Proguanil/ or Phenformin/<br>8. (biguanid* or metformin* or buformin* or chlorhexidin* or chlorguanid* or phenformin*).ti,ab,kw.<br>9. 7 or 8<br>10. 6 or 9<br>11. Blood Glucose/ or Diabetes, Gestational/ or Glucose Tolerance Test/ or Insulin/ or Insulin Resistance/ or Obesity/ or Overweight/<br>12. (gestational weight gain or glucose* or glycemic* or insulin resistan*).ti,ab,kw.<br>13. ((obes* or overweight or weight gain) adj3 (women or matern* or pregnan*)).ti,ab,kw.<br>14. 11 or 12 or 13<br>15. "Clinical Trials as Topic"/ or Controlled Clinical Trial/ or Randomized Controlled Trial/<br>16. (trial or randomiz* or RCT or randomis* or control group* or two arm* or (two adj2 arm) or quasiexperiment* or (quasi* adj2 experiment*) or (matched adj2 (cohort* or control*))).ti,ab.<br>17. randomi*.pt.<br>18. (intervention or assigned* or compar*).ti,ab.<br>19. 15 or 16 or 17 or 18<br>20. 3 and 10 and 14 and 19<br>21. 20 not (Animals/ not (Animals/ and Humans/)) |
| **Embase**<br><br>#1. antenatal:ti,ab OR ((ante* NEAR/2 natal):ti,ab) OR childbearing:ti,ab OR ((child NEAR/2 bearing):ti,ab) OR gestation*:ti,ab OR 'family planning services':ti,ab OR gravid*:ti,ab OR interconcept*:ti,ab OR intergestation*:ti,ab OR internatal:ti,ab OR matern*:ti,ab OR periconcept*:ti,ab OR preconcept*:ti,ab OR pregestation*:ti,ab OR prenatal*:ti,ab OR prepregn*:ti,ab OR (((inter* OR pre* OR peri*)<br>NEAR/2 (concept* OR gestation* OR natal OR pregnan*)):ti,ab)<br><br>#2. 'pregnancy'/exp OR 'prepregnancy care'/exp |

#3. #1 OR #2

#4. 'behavior therapy'/exp OR 'cognitive behavioral therapy'/exp OR 'diet'/exp OR 'exercise'/exp OR 'health behavior'/exp OR 'health education'/exp OR 'health promotion'/exp OR 'lifestyle'/exp OR 'body weight loss'/exp

#5 (((behav* NEAR/2 (cognit* OR manag* OR modif* OR therap*)):ti,ab) OR cbt:ti,ab OR diet*:ti,ab OR ((health* NEAR/2 (behav* OR eat* OR educat* OR food* OR promotion)):ti,ab) OR lifestyle:ti,ab OR ((life NEAR/2 style):ti,ab) OR nutrition:ti,ab OR physical:ti,ab) AND activit*:ti,ab OR ((weight NEAR/2 (loss OR manag* OR reduction OR retention)):ti,ab)

#6. #4 OR #5

#7. 'biguanide'/exp OR 'metformin'/exp OR 'buformin'/exp OR 'chlorhexidine'/exp OR 'proguanil'/exp OR 'phenformin'/exp

#8. biguanid*:ti,ab OR metformin*:ti,ab OR buformin*:ti,ab OR chlorhexidin*:ti,ab OR chlorguanid*:ti,ab OR phenformin*:ti,ab

#9. #7 OR #8

#10. #6 OR #9

#11. 'glucose blood level'/exp OR 'pregnancy diabetes mellitus'/exp OR 'glucose tolerance test'/exp OR 'insulin'/exp OR 'insulin resistance'/exp OR 'obesity'/exp

#12. 'gestational weight gain':ti,ab OR glucose*:ti,ab OR glycemic*:ti,ab OR glycaemic*:ti,ab OR ((insulin* NEAR/2 resist*):ti,ab)

#13. ((obes* OR overweight OR 'weight gain') NEAR/3 (women OR matern* OR pregnan*)):ti,ab

#14. #11 OR #12 OR #13

#15. 'crossover procedure':de OR 'double-blind procedure':de OR 'randomized controlled trial':de OR 'single-blind procedure':de OR random*:de,ab,ti OR factorial*:de,ab,ti OR crossover*:de,ab,ti OR ((cross NEXT/1 over*):de,ab,ti) OR placebo*:de,ab,ti OR ((doubl* NEAR/1 blind*):de,ab,ti) OR ((singl* NEAR/1 blind*):de,ab,ti) OR assign*:de,ab,ti OR allocat*:de,ab,ti OR volunteer*:de,ab,ti

#16. #3 AND #10 AND #14 AND #15 AND [humans]/lim

**Pubmed**

#1
("Pregnancy"[Mesh] OR "Gravidity"[Mesh] OR "Preconception Care"[Mesh]) OR (gravid*[Title/Abstract] OR pregnan*[Title/Abstract] OR childbearing[Title/Abstract] OR child bearing[Title/Abstract] OR gestation*[Title/Abstract] OR matern*[Title/Abstract] OR preconcept*[Title/Abstract] OR prenatal[Title/Abstract] OR antenatal[Title/Abstract] OR pre-concept*[Title/Abstract] OR "pre concept*"[Title/Abstract] OR prepregnan*[Title/Abstract] OR pre-pregnan*[Title/Abstract] OR "pre-pregnan*"[Title/Abstract] OR pregestation*[Title/Abstract] OR pre-gestation*[Title/Abstract] OR "pre

gestation*"[Title/Abstract] OR periconcept*[Title/Abstract] OR peri-concept*[Title/Abstract] OR "peri concept*"[Title/Abstract] OR interconcept*[Title/Abstract] OR inter-concept*[Title/Abstract] OR "inter concept*"[Title/Abstract] OR interpregnan*[Title/Abstract] OR inter-pregnan*[Title/Abstract] OR "inter pregnan*"[Title/Abstract] OR intergestation*[Title/Abstract] OR inter-gestation*[Title/Abstract] OR "inter gestation*"[Title/Abstract] OR internatal[Title/Abstract] OR "family planning services"[Title/Abstract] OR "family-planning services"[Title/Abstract] OR ante-natal[Title/Abstract] OR pre-natal[Title/Abstract])

#2
(behavior therap*[Title/Abstract] OR behaviour therap*[Title/Abstract] OR behavioral therap*[Title/Abstract] OR behavioural therap*[Title/Abstract] OR behavior modif*[Title/Abstract] OR behaviour modif*[Title/Abstract] OR behavioral modif*[Title/Abstract] OR behavioural modif*[Title/Abstract] OR behavior manage*[Title/Abstract] OR behaviour manage*[Title/Abstract] OR behavioral manage*[Title/Abstract] OR behavioural manage*[Title/Abstract] OR cbt[Title/Abstract] OR diet*[Title/Abstract] OR health behavior*[Title/Abstract] OR health education[Title/Abstract] OR health promotion[Title/Abstract] OR healthy eat*[Title/Abstract] OR healthy food[Title/Abstract] OR life style[Title/Abstract] OR lifestyle[Title/Abstract] OR life-style[Title/Abstract] OR nutrition[Title/Abstract] OR physical activit*[Title/Abstract] OR weight loss[Title/Abstract] OR weight management[Title/Abstract] OR weight reduction[Title/Abstract] OR weight retention[Title/Abstract]) OR ("Behavior Therapy"[Mesh] OR "Cognitive Behavioral Therapy"[Mesh] OR "Diet"[Mesh] OR "Exercise"[mesh] OR "Health Behavior"[mesh] OR "Health Education"[Mesh] OR "Health Promotion"[Mesh] OR "Life Style"[Mesh] OR "Weight Loss"[Mesh])

#3
("Biguanides"[Mesh] OR "Metformin"[Mesh] OR "Buformin"[Mesh] OR "Chlorhexidine"[Mesh] OR "Proguanil"[Mesh] OR "Phenformin"[Mesh]) OR (biguanid*[Title/Abstract] OR metformin*[Title/Abstract] OR buformin*[Title/Abstract] OR chlorhexidin*[Title/Abstract] OR chlorguanid*[Title/Abstract] OR phenformin*[Title/Abstract])

#4
#2 OR #3

#5
("Blood Glucose"[mesh] OR "Diabetes, Gestational"[Mesh] OR "Insulin"[mesh] OR "Insulin Resistance" OR "Glucose Tolerance Test"[mesh] OR "Obesity"[mesh] OR "Overweight"[mesh]) OR ("gestational weight gain"[Title/Abstract] OR "glucose intolerance" [Title/Abstract] OR glucose*[Title/Abstract] OR glycemic*[Title/Abstract] OR “insulin resistan*”[Title/Abstract] OR obesity[Title/Abstract])

#6
“Clinical Trials as Topic"[Mesh] OR "Randomized Controlled Trial"[Publication Type] OR trial[Title/Abstract] OR randomiz*[Title/Abstract] OR RCT[Title/Abstract] OR randomis*[Title/Abstract] OR "control group*"[Title/Abstract] OR “two-arm*”[Title/Abstract] OR "two arm"[Title/Abstract] OR “quasi-experiment*”[Title/Abstract] OR "matched cohort*"[Title/Abstract] OR "matched control*"[Title/Abstract] OR intervention[Title/Abstract] OR assigned*[Title/Abstract] OR compar*[Title/Abstract]

#7
#1 AND #4 AND #5 AND #6

Supplementary Table 2: Inclusion criteria

| **Population** | **Intervention** | **Outcome** | **Limits** |
|---|---|---|---|
| All women (of childbearing age) | Interventions:<br>● Diet<br>● Exercise<br>● Behavioral<br>● Lifestyle<br>● Combined (diet, behavioral, exercise)<br>● Metformin<br>● Supplementation<br><br>Control:<br>● Usual care<br>● Placebo<br>● Minimal intervention (e.g. not more than 1 session a year | Primary<br>Gestational diabetes<br><br>Secondary<br>Gestational weight gain | Randomised or non randomized controlled trials<br><br>Language: English<br><br>Years of publication: all years |

Supplementary Table 3: Definition of participant characteristics

| **Category** | **Definition** |
|---|---|
| Gestational age | ● Preconception<br>● 1st trimester: ≤12 gestational week<br>● Early 2nd trimester: 13-17 weeks<br>● Late 2nd trimester: ≥18-26 weeks<br>(if it was a range, the highest number of the range was considered) |
| Educational status | ● Tertiary education: if ≥50% of the participants had attended tertiary education.<br>● Not attended tertiary education: if <50% of the participants had attended tertiary education. |
| Employment | ● Employed: if ≥50% of the participants were employed.<br>● Unemployed: if <50% of the participants were employed. |
| Ethnicity | ● White: ≥80% of the participants were of European and other Caucasian origins.<br>● Non-white: if <80 were white.<br>● Mixed: a combination of White and non-White where neither was >80% for each group |
| BMI (based on inclusion criteria) | ● Normal weight: if only normal-weight participants were recruited.<br>● Overweight obese: if overweight/obese only participants were recruited. |
| Parity | ● Nulliparous: if all participants were nulliparous<br>● Not nulliparous: If none of the participants were nulliparous<br>● Mixed: A mixture of nulliparous and not nulliparous |
| Hypertension (based on inclusion criteria) | ● With: if all participants had hypertension. |

| | |
|---|---|
| | • Without: if all participants were free of hypertension. |
| Prediabetes (based on inclusion criteria) | • With: if all participants had prediabetes.<br>• Without: if all participants were free of prediabetes. |
| Polycystic ovary syndrome (PCOS) (based on inclusion criteria) | • With: if all participants had PCOS.<br>• Without: if all participants were free of PCOS. |
| Cardiovascular disease (CVD) (based on inclusion criteria) | • With: if all participants had CVD<br>• Without: if all participants were free of CVD |
| Smoking (based on inclusion criteria) | • With: if all participants had a history of smoking<br>• Without: if all participants were free of history of smoking |
| History of macrosomia (based on inclusion criteria) | • With: if all participants had a history of macrosomia<br>• Without: if all participants were free of history of macrosomia |
| History of gestational diabetes (GDM)(based on inclusion criteria) | • With: if all participants had a history of GDM<br>• Without: if all participants were free of history of GDM |
| History of hypertensive disorders during pregnancy (HDP) (based on inclusion criteria) | • With: if all participants had a history of HDP<br>• Without: if all participants were free of history of HDP |
| History of stillbirth (based on inclusion criteria) | • With: if all participants had a history of stillbirth<br>• Without: if all participants were free of history of stillbirth |

Supplementary Table 4: Participant characteristics of included studies

| **Author, year** | **Trimester** | **BMI** | **Tertiary educated** | **Employment** | **Ethnicity** | **Hypertension** | **Hyperlipidemia** | **Prediabetes** | **Parity** | **PCOS** | **Stillbirth** | **Familyhistoryofdiabetes** | **PriorLGAormacrosomiainfant** | **PasthistoryofGDM** | **PasthistoryofHDP** | **Historyofcardiovasculardiseas** | **Smoking** |
|---|---|---|---|---|---|---|---|---|---|---|---|---|---|---|---|---|---|
| Abdel-Aziz, 2018 | 1st | All BMIs | No | No | NR | NR | NR | NR | Nulliparous | NR | No | NR | NR | NR | NR | NR | NR |
| Adb El Hameed, 2011 | Preconception | All BMIs | NR | NR | NR | NR | NR | NR | NR | Yes | NR | NR | NR | NR | NR | NR | NR |
| Ainuddin, 2015 | Preconception | All BMIs | NR | NR | NR | NR | NR | NR | Mixed | Yes | NR | NR | NR | NR | NR | NR | NR |
| Alamolhoda, 2019 | 1st | All BMIs | No | No | NR | No | NR | No | NR | NR | No | No | No | No | No | No | No |
| Alwattar, 2018 | Early 2nd | All BMIs | NR | NR | Mixed | NR | NR | No | Mixed | NR | NR | NR | NR | No | NR | NR | NR |
| Assaf, 2017 | 1st | All BMIs | Yes | NR | Mixed | NR | NR | NR | Mixed | NR | NR | NR | NR | NR | NR | NR | NR |
| Barakat, 2013 | 1st | All BMIs | No | Yes | NR | No | NR | No | NR | NR | NR | NR | NR | NR | NR | NR | NR |
| Barakat, 2012 | 1st | All BMIs | No | Yes | NR | NR | NR | NR | Mixed | NR | NR | NR | NR | NR | NR | NR | NR |
| Barakat, 2019 | 1st | All BMIs | No | NR | NR | NR | NR | No | Mixed | NR | NR | NR | NR | No | NR | NR | NR |
| Barakat, 2014 | 1st | All BMIs | No | NR | NR | NR | NR | NR | Mixed | NR | NR | NR | NR | NR | No | No | NR |
| Basu, 2021 | Late 2nd | Obese | NR | NR | Non-Caucasians | No | NR | NR | Mixed | NR | NR | NR | NR | NR | NR | NR | NR |
| Bogaerts, 2013 | Early 2nd | Obese | No | Yes | Mixed | NR | NR | No | Mixed | NR | NR | NR | NR | NR | NR | NR | NR |
| Bruno, 2016 | 1st | Overweight/obese | No | Yes | Caucasians | No | NR | NR | Mixed | NR | NR | NR | NR | No | NR | NR | No |
| Buckingham-Schutt, 2019 | Early 2nd | All BMIs | NR | NR | NR | No | NR | NR | NR | NR | NR | NR | NR | No | No | No | NR |
| Cahill 2018, 2018 | Early 2nd | Overweight/obese | No | NR | NR | NR | NR | No | Mixed | NR | NR | NR | No | No | NR | NR | No |
| Callaway, 2010 | 1st | Obese | NR | NR | NR | NR | NR | NR | NR | NR | NR | NR | NR | NR | NR | NR | NR |
| Callaway, 2019 | Late 2nd | Overweight/obese | NR | NR | Caucasians | NR | NR | NR | Mixed | NR | NR | NR | NR | NR | NR | NR | NR |
| Celentano, 2010 | 1st | Obese | NR | NR | NR | NR | NR | NR | Mixed | NR | NR | NR | NR | NR | NR | NR | NR |
| Chan R, 2018 | 1st | All BMIs | Yes | Yes | Non-Caucasians | NR | NR | No | Mixed | NR | NR | NR | NR | NR | NR | NR | No |
| Chiswick, 2008 | Early 2nd | Overweight/obese | NR | NR | Caucasians | NR | NR | No | NR | NR | NR | NR | NR | No | No | NR | NR |

| | | | | | | | | | | | | | | | | | |
|---|---|---|---|---|---|---|---|---|---|---|---|---|---|---|---|---|---|
| Cordero, 2015 | 1st | All BMIs | NR | Yes | NR | NR | NR | NR | Mixed | NR | NR | NR | NR | NR | NR | NR | NR |
| D'Anna, 2013 | Early 2nd | Obese | NR | NR | Caucasians | NR | NR | No | Mixed | NR | NR | Yes | NR | No | NR | NR | NR |
| D'Anna, 2015 | Early 2nd | Nomal and overweight | NR | NR | NR | No | NR | No | Mixed | NR | NR | NR | NR | No | NR | NR | NR |
| DaSilva, 2017 | Late 2nd | All BMIs | NR | Yes | Mixed | No | NR | No | NR | NR | NR | NR | NR | NR | NR | No | NR |
| Deng, 2022 | Early 2nd | All BMIs | No | NR | NR | NR | NR | NR | NR | NR | NR | NR | NR | NR | NR | NR | NR |
| Ding , 2021 | 1st | Overweight/obese | NR | Yes | NR | NR | NR | No | Mixed | No | NR | No | No | No | NR | NR | NR |
| Dodd, 2018 | Late 2nd | All BMIs | NR | NR | Caucasians | NR | NR | No | Mixed | NR | NR | NR | NR | NR | NR | NR | NR |
| Dodd , 2019 | Late 2nd | Overweight/obese | NR | NR | Mixed | NR | NR | No | Mixed | NR | NR | NR | NR | NR | NR | NR | NR |
| Epel, 2019 | Late 2nd | Overweight/obese | No | NR | Non-Caucasians | No | NR | No | NR | No | NR | NR | NR | NR | NR | No | NR |
| Eslami , 2018 | Late 2nd | Overweight/obese | No | No | NR | NR | NR | NR | Mixed | NR | NR | NR | NR | No | NR | NR | NR |
| Farren, 2017 | Early 2nd | All BMIs | NR | NR | Mixed | NR | NR | No | Mixed | NR | NR | Yes | NR | NR | NR | NR | NR |
| Garmendia, 2020 | Early 2nd | All BMIs | No | NR | NR | NR | NR | NR | Mixed | NR | NR | NR | NR | NR | NR | NR | NR |
| Glueck, 2002 | NR | All BMIs | NR | NR | Caucasians | NR | NR | No | NR | Yes | NR | NR | NR | NR | NR | NR | NR |
| Glueck, 2022 | Preconception | Overweight/obese | NR | NR | NR | NR | NR | NR | Mixed | Yes | NR | NR | NR | NR | NR | NR | NR |
| Gonzalez-Plaza , 2022 | Early 2nd | Obese | No | Yes | NR | No | NR | No | Mixed | NR | NR | NR | NR | NR | NR | No | NR |
| Gray-Donald, 2000 | Late 2nd | All BMIs | NR | NR | NR | NR | NR | No | Mixed | NR | NR | NR | NR | NR | NR | NR | NR |
| Gregory, 2016 | Early 2nd | Obese | NR | NR | Non-Caucasians | NR | NR | No | Mixed | NR | NR | NR | NR | NR | NR | No | NR |
| Hajian, 2020 | Late 2nd | Overweight | No | NR | NR | NR | NR | NR | Mixed | NR | NR | NR | NR | NR | NR | NR | NR |
| Harrison, 2012 | Early 2nd | Overweight/obese | Yes | Yes | NR | NR | NR | No | NR | NR | NR | NR | NR | NR | NR | NR | NR |
| Herring, 2016 | 1st | Overweight/obese | No | NR | Non-Caucasians | NR | NR | NR | Mixed | NR | NR | NR | NR | NR | NR | NR | NR |
| Hui, 2004 | Late 2nd | All BMIs | No | NR | Mixed | NR | NR | No | NR | NR | NR | NR | NR | NR | NR | NR | NR |
| Hui, 2012 | Late 2nd | All BMIs | NR | NR | Mixed | NR | NR | No | NR | NR | NR | NR | NR | NR | NR | NR | NR |
| Hui, 2014 | Late 2nd | All BMIs | NR | NR | NR | NR | NR | NR | Mixed | NR | NR | NR | NR | NR | NR | NR | NR |

| | | | | | | | | | | | | | | | | | |
|---|---|---|---|---|---|---|---|---|---|---|---|---|---|---|---|---|---|
| Jamal, 2012 | 1st | All BMIs | NR | NR | NR | No | NR | No | Mixed | Yes | NR | NR | NR | NR | NR | NR | NR |
| Janumalal, 2020 | Early 2nd | Overweight/obese | NR | NR | Mixed | NR | NR | NR | NR | NR | NR | NR | NR | NR | NR | NR | NR |
| Jing, 2015 | 1st | All BMIs | NR | NR | NR | NR | NR | No | NR | NR | NR | NR | NR | NR | NR | NR | NR |
| Jovanovic-Peterson, 1997 | NR | All BMIs | NR | NR | Non-Caucasians | No | NR | NR | NR | NR | NR | NR | NR | NR | NR | NR | NR |
| Kennelly, 2018 | Early 2nd | Overweight/obese | Yes | NR | Caucasians | NR | NR | NR | Mixed | NR | NR | NR | NR | NR | NR | NR | NR |
| Khattab, 2011 | NR | Overweight/obese | NR | NR | NR | NR | NR | NR | Mixed | Yes | NR | NR | NR | NR | NR | NR | NR |
| Ko, 2012 | Late 2nd | All BMIs | NR | NR | Mixed | NR | NR | NR | NR | NR | NR | NR | NR | NR | NR | NR | NR |
| Koivusalo 2016, Huvinen 2018,Rono 2018, Huvinen 2022, Valkama 2018, Grotenfelt 2019 , 2018 | Late 2nd | All BMIs | No | NR | NR | NR | NR | NR | Mixed | NR | NR | NR | NR | NR | NR | NR | NR |
| Kong, 2014 | Early 2nd | Overweight/obese | NR | Yes | NR | No | NR | No | NR | NR | NR | NR | NR | No | NR | No | No |
| Korpi-Hyovalti, 2011 | 1st | All BMIs | No | Yes | NR | NR | NR | NR | Mixed | NR | NR | NR | NR | NR | NR | NR | NR |
| Kunath 2019, Gunther 2022, Hoffman 2021, 2019 | 1st | All BMIs | No | NR | NR | NR | NR | No | Mixed | NR | NR | NR | NR | NR | NR | NR | NR |
| LeBlan, 2020&2021 | Preconception | Overweight/obese | Yes | NR | Caucasians | NR | NR | NR | NR | NR | NR | NR | NR | NR | NR | NR | NR |
| Li, 2021 | NR | All BMIs | NR | NR | NR | NR | NR | NR | Mixed | NR | NR | NR | NR | NR | NR | NR | NR |
| Lin, 2020 | 1st | All BMIs | NR | NR | NR | NR | NR | NR | Mixed | NR | NR | NR | NR | NR | NR | NR | NR |
| Lindsay, 2014 | Late 2nd | All BMIs | No | NR | Caucasians | NR | NR | No | Mixed | NR | NR | NR | NR | No | NR | NR | NR |
| Liu, 2015 | 1st | Overweight/obese | NR | NR | NR | No | NR | No | NR | Yes | NR | NR | NR | NR | NR | NR | NR |
| Liu, 2021 | Early 2nd | Overweight/obese | Yes | Yes | Mixed | NR | NR | NR | Mixed | NR | NR | NR | NR | NR | NR | NR | NR |
| Liu, 2015 | Early 2nd | Overweight/obese | No | Yes | NR | No | NR | No | NR | NR | NR | NR | NR | NR | NR | No | NR |
| Lovvik, 2019 | 1st | All BMIs | Yes | Yes | Caucasians | NR | NR | NR | Mixed | Yes | NR | NR | NR | NR | NR | NR | NR |
| Luoto, 2010 | 1st | All BMIs | Yes | NR | NR | NR | NR | NR | Mixed | NR | NR | NR | NR | NR | NR | NR | NR |
| Luoto, 2010 | 1st | All BMIs | Yes | NR | NR | NR | NR | NR | Mixed | NR | NR | NR | NR | NR | NR | NR | NR |

| | | | | | | | | | | | | | | | | | |
|---|---|---|---|---|---|---|---|---|---|---|---|---|---|---|---|---|---|
| Luoto and Kolu, 2011 | 1st | All BMIs | Yes | NR | NR | NR | NR | NR | Mixed | NR | NR | NR | NR | NR | NR | NR | NR |
| Matarrelli , 2013 | Early 2nd | All BMIs | NR | NR | NR | NR | NR | Yes | Mixed | NR | NR | NR | NR | NR | NR | NR | NR |
| McCarthy EA, 2016 | Late 2nd | Overweigh t/obese | Yes | NR | NR | NR | NR | NR | Mixed | NR | NR | NR | NR | NR | NR | NR | NR |
| Mohsenzadeh -ledari F, 2020 | Late 2nd | All BMIs | No | No | NR | NR | NR | NR | Mixed | NR | NR | NR | NR | NR | NR | NR | NR |
| Motahari-Tabari N, 2021, 2021 | Early 2nd | Overweigh t/obese | Yes | No | NR | NR | NR | No | Mixed | NR | NR | NR | NR | No | NR | NR | NR |
| Oostdam, 2012 | Early 2nd | Obsese | No | Yes | Mixed | No | NR | NR | Mixed | NR | NR | NR | NR | NR | NR | NR | NR |
| Opie , 2016 | Late 2nd | All BMIs | NR | NR | NR | NR | NR | No | Mixed | NR | NR | NR | NR | NR | NR | No | No |
| Parat , 2019 | Late 2nd | Overweigh t/obese | Yes | NR | NR | NR | NR | No | Mixed | NR | NR | NR | NR | NR | NR | NR | NR |
| Peccei, 2017 | 1st | Overweigh t/obese | No | NR | Mixed | NR | NR | NR | NR | NR | NR | NR | NR | NR | NR | NR | NR |
| Pelaez, , 2019 | Early 2nd | Overweigh t/obese | NR | NR | NR | NR | NR | NR | NR | NR | NR | NR | NR | NR | NR | NR | NR |
| Pellonper¨a, 2019 | 1st | Overweigh t/obese | Yes | NR | Cauca sians | NR | NR | NR | Mixed | NR | NR | NR | NR | NR | NR | NR | NR |
| Petrella , 2014 | Early 2nd | Overweigh t/obese | No | Yes | Mixed | No | NR | No | Mixed | NR | NR | NR | NR | No | NR | NR | NR |
| Phelan , 2011 | Early 2nd | All BMIs | Yes | NR | Mixed | NR | NR | NR | Mixed | NR | NR | NR | NR | NR | NR | NR | NR |
| Phelan , 2018 | Early 2nd | Overweigh t/obese | Yes | NR | Mixed | NR | NR | No | Mixed | NR | NR | NR | NR | NR | NR | NR | NR |
| Phillips, 2019 | Early 2nd | Overweigh t/obese | No | NR | Cauca sians | NR | NR | NR | Mixed | NR | NR | NR | NR | NR | NR | NR | NR |
| Polley , 2002 | Late 2nd | All BMIs | Yes | NR | Mixed | NR | NR | NR | NR | NR | NR | NR | NR | NR | NR | NR | NR |
| Poston 2015, Mills 2019, Peacock 2020, 2015 | Late 2nd | Obese | NR | NR | Mixed | No | NR | No | Mixed | NR | NR | NR | NR | NR | NR | NR | NR |
| Price , 2011 | Early 2nd | All BMIs | NR | NR | Mixed | No | NR | NR | Mixed | NR | NR | NR | NR | NR | NR | NR | NR |
| Quinlivan , 2011 | NR | Overweigh t obese | NR | NR | Mixed | NR | NR | NR | Mixed | NR | NR | NR | NR | NR | NR | NR | NR |
| Rauh , 2013 | Late 2nd | All BMIs | Yes | Yes | NR | NR | NR | NR | Mixed | NR | NR | NR | NR | NR | NR | NR | NR |
| Renault , 2014 | Early 2nd | Obese | NR | NR | Cauca sians | NR | NR | No | Mixed | NR | NR | NR | NR | NR | NR | NR | NR |
| Ruiz , 2013 | 1st | All BMIs | No | Yes | NR | NR | NR | NR | NR | NR | NR | NR | NR | NR | NR | NR | NR |
| Sagedal, 2016&17 | Late 2nd | All BMIs | No | Yes | NR | NR | NR | No | Nullip arous | NR | NR | NR | NR | NR | NR | NR | NR |

| | | | | | | | | | | | | | | | | | |
|---|---|---|---|---|---|---|---|---|---|---|---|---|---|---|---|---|---|
| Sahariah , 2016 | Preconception | All BMIs | No | No | Non-Caucasians | No | NR | NR | Mixed | NR | NR | NR | NR | NR | NR | NR | NR |
| Sales , 2018 | Late 2nd | Obese | No | Yes | Caucasians | NR | NR | NR | NR | NR | NR | NR | NR | NR | NR | NR | NR |
| Santamaria, 2016 | Early 2nd | Overweight | NR | NR | Caucasians | NR | NR | No | Mixed | NR | NR | NR | NR | No | NR | NR | NR |
| Seneviratne, 2015 | Late 2nd | Overweight/obese | NR | Yes | Mixed | NR | NR | NR | Mixed | NR | NR | NR | NR | NR | NR | NR | No |
| Shirazian, 2010 | Early 2nd | Obese | NR | NR | Non-Caucasians | No | NR | No | Mixed | NR | NR | NR | NR | NR | NR | NR | NR |
| Shirazian, 2016 | Early 2nd | Obese | NR | NR | Non-Caucasians | No | NR | No | Mixed | NR | NR | NR | NR | NR | NR | NR | NR |
| Simmons, 2017 | Late 2nd | Obese | Yes | NR | Caucasians | NR | NR | No | Mixed | NR | NR | NR | NR | NR | NR | NR | NR |
| Stafne, 2012 | Late 2nd | All BMIs | NR | NR | NR | NR | NR | NR | Mixed | NR | NR | NR | NR | NR | NR | NR | NR |
| Sun, 2020 | Preconception | All BMIs | No | Yes | Non-Caucasians | No | NR | No | NR | NR | NR | NR | NR | NR | NR | NR | NR |
| Sun, 2016 | 1st | Overweight/obese | Yes | NR | NR | NR | NR | NR | NR | NR | NR | NR | NR | NR | NR | NR | NR |
| Syngelaki, 2016 | Early 2nd | Obese | NR | NR | Mixed | NR | NR | NR | Mixed | NR | NR | NR | NR | No | NR | NR | NR |
| Thornton, 2009 | Late 2nd | Obese | NR | NR | Mixed | No | NR | NR | Mixed | NR | NR | NR | NR | NR | NR | NR | NR |
| Tomić , 2013 | 1st | All BMIs | No | NR | NR | No | NR | No | Mixed | NR | NR | NR | NR | NR | NR | No | NR |
| Trak-Fellermeier 2019, Haslam 2020, 2019 | Early 2nd | Overweight/obese | Yes | NR | Mixed | NR | NR | No | Mixed | NR | NR | NR | NR | NR | NR | NR | No |
| Valdés, 2018 | Early 2nd | All BMIs | NR | NR | NR | NR | NR | No | Mixed | NR | NR | NR | NR | NR | NR | NR | NR |
| Van Horn, , 2018 | Early 2nd | Overweight/obese | Yes | NR | Mixed | NR | NR | No | Mixed | NR | NR | NR | NR | NR | NR | NR | No |
| VANKY, 2010 | Early 2nd | All BMIs | NR | NR | NR | NR | NR | NR | Mixed | Yes | NR | NR | NR | NR | NR | NR | NR |
| Vesco , 2014 | Late 2nd | Obese | Yes | NR | Caucasians | NR | NR | No | NR | NR | NR | NR | NR | NR | NR | NR | NR |
| Vinter 2011 Vinter 2014, 2014 | Early 2nd | Obese | Yes | Yes | NR | No | NR | No | Mixed | NR | NR | NR | NR | No | NR | No | NR |
| Vitale , 2021 | Early 2nd | Overweight | NR | NR | NR | NR | NR | No | Mixed | NR | NR | NR | NR | No | NR | NR | NR |
| Walsh , 2012 | Late 2nd | Normal weight | NR | NR | NR | NR | NR | NR | Not nulliparous | NR | NR | NR | Yes | No | NR | NR | NR |

| | | | | | | | | | | | | | | | | | |
|---|---|---|---|---|---|---|---|---|---|---|---|---|---|---|---|---|---|
| Wang , 2015 | 1st | All BMIs | NR | NR | NR | NR | NR | NR | NR | NR | NR | NR | NR | NR | NR | NR | NR |
| Wang , 2017 | Early 2nd | Overweight | Yes | NR | NR | No | NR | No | Mixed | NR | NR | NR | NR | NR | NR | No | No |
| Wickens, 2017 | Early 2nd | All BMIs | NR | NR | Mixed | NR | NR | NR | Mixed | NR | NR | NR | NR | NR | NR | NR | NR |
| Wolff , 2008 | Early 2nd | Obese | NR | NR | Caucasians | NR | NR | NR | NR | NR | NR | NR | NR | NR | NR | NR | No |
| Xu , 2022 | Late 2nd | All BMIs | NR | NR | NR | NR | NR | NR | Mixed | NR | NR | NR | NR | NR | NR | NR | NR |
| Zhang , 2015 | 1st | All BMIs | NR | NR | NR | NR | NR | NR | NR | NR | NR | NR | NR | NR | NR | NR | NR |
| Zhang , 2019 | Early 2nd | Overweight/obese | NR | NR | NR | No | NR | No | NR | NR | NR | NR | NR | NR | NR | No | NR |
| Zhao , 2022 | 1st | All BMIs | No | Yes | NR | NR | NR | NR | Mixed | No | NR | NR | NR | NR | NR | NR | NR |

BMI: Body mass index, PCOS: polycystic ovary syndrome; GDM: Gestational diabetes; HDP: Hypertensive disorder during pregnancy; CVD: cardiovascular disease; NR: Not reported; Yes: With condition; No: Without condition;

Supplementary Table 5: Risk of bias summary

Supplementary Table 5a: Risk of bias of randomised controlled trials

| Author, year | Randomization process | Deviations from intended interventions | Missing outcome data | Measurement of the outcome | Selection of the reported result | Overall bias |
|---|---|---|---|---|---|---|
| Abdel Aziz, 2018 | Some concerns | Low | Low | Low | Low | Some concerns |
| Alamodhoda, 2019 | Some concerns | Low | Low | Low | Low | Some concerns |
| Alwatter, 2018 | Some concerns | High | Low | Low | Some concerns | High |
| Assaf-Balut, 2017 | High | High | High | Low | Low | High |
| Barakat, 2013 | Low | Low | Low | Low | Low | Some concerns |
| Barakat, 2014 | Some concerns | High | Low | Low | Low | High |
| Barakat, 2019 | Low | Some concerns | Low | Low | Low | Some concerns |
| Basu, 2021 | Some concerns | High | Low | Low | Low | Some concerns |
| Bogaerts 2013 | Low | Low | Low | Low | Low | Low |
| Barakat, 2012 | Some concerns | High | Low | Low | Low | High |
| Bruno 2016 | Low | Low | Low | Low | Low | Low |
| Buckingham-Schutt 2019 | Some concerns | High | Low | Low | Low | High |
| Cahil 2018 | High | Low | Low | Low | Low | High |
| Callaway 2010 | Some concerns | High | High | Low | Low | High |
| Celentano 2020 | Some concerns | Low | Low | Low | Low | Some concerns |
| Chan R, 2018 | Low | Low | Low | Low | Low | Low |
| Chiswick 2008 | Low | Low | Low | Low | Low | Low |
| Cordero, 2015 | Some concerns | Low | Low | Low | Some concerns | Some concerns |
| D’Anna 2015 | Some concerns | Some concerns | Low | Low | Low | Some concerns |
| D'Anna, 2013 | Low | Low | Low | Low | Low | Low |
| Da Silva 2017 | Low | High | High | Low | Low | High |
| Deng, 2022 | High | Low | Low | Low | Low | High |
| Ding 2021 | Low | Low | Low | Some concerns | Low | Some concerns |
| Dodd, 2018 | Low | Low | Low | Low | Low | Low |
| Dodd, 2019 | Low | Low | Low | Low | Low | Low |
| Eslami, 2018 | Some concerns | Some concerns | Low | Some concerns | Low | Some concerns |
| Farren 2017 | Low | High | Low | Low | Low | High |
| Gonzalez-Plaza, 2022 | Low | Some concerns | Low | Low | Some concerns | Some concerns |
| Guelfi, 2016 | Low | Low | Low | Low | Some concerns | Some concerns |
| Harrison, 2016 | Low | Low | Low | Low | Some concerns | Some concerns |
| Herring, 2016 | Low | Low | Low | Low | Some concerns | Some concerns |

| | | | | | | |
|---|---|---|---|---|---|---|
| Hui 2006 | Some concerns | Some concerns | Low | Low | Some concerns | Some concerns |
| Hui 2012 | Low | Some concerns | Low | Low | Low | Some concerns |
| Hui 2014 | Low | Some concerns | Low | Low | Low | Some concerns |
| Jamal 2012 | Low | Some concerns | Low | Low | Low | Some concerns |
| Janumala 2020 | Some concerns | High | Low | Low | Some concerns | High |
| Jing 2015 | Low | Some concerns | Low | Low | Low | Some concerns |
| Jovanovic-Peterson 1997 | Some concerns | Some concerns | Low | Low | Some concerns | Some concerns |
| Kennelly 2018 | Low | Some concerns | Low | Low | Low | Some concerns |
| Ko 2014 | Some concerns | Some concerns | Low | Low | Some concerns | Some concerns |
| Kong, 2013 | Low | Low | Low | Some concerns | Some concerns | Some concerns |
| Korpi-Hyovalti | Low | Some concerns | Low | Low | Some concerns | Some concerns |
| LeBlanc, 2020 | Low | Some concerns | Low | Low | Some concerns | Some concerns |
| Li, 2021 | Low | Some concerns | Low | Low | Some concerns | Some concerns |
| Lin, 2020 | Low | Some concerns | Low | Low | Some concerns | Some concerns |
| Lindsay 2014 | Some concerns | High | Low | Low | Low | High |
| Liu, 2021 | Low | Some concerns | Some concerns | Low | Low | Some concerns |
| Luoto, 2010 | Low | Low | High | Some concerns | Low | Some concerns |
| Luoto, 2010 | Low | Low | High | Some concerns | Low | Some concerns |
| Matarrelli 2013 | Low | Low | Low | Low | Low | Low |
| McCarthy | Low | Low | Low | Low | Low | Low |
| Mohsenzadeh, 2020 | Some concerns | Low | Low | Low | Some concerns | Low |
| Motahari-Tabari, 2021 | Low | Some concerns | Low | Low | Low | Some concerns |
| Ostdam | Low | High | High | Low | Low | High |
| Parat 2018 | Low | High | Low | Low | Low | High |
| Peccei 2017 | Some concerns | High | Low | Low | Low | High |
| Pelaez 2019 | Some concerns | Low | Low | Low | Low | Some concerns |
| Pellonpera 2019 | High | Low | Low | Low | Low | High |
| Petrella 2014 | Some concerns | High | Low | Low | Low | High |
| Phelan 2011 | Low | Some concerns | Low | Low | Low | Some concerns |
| Phelan 2018 | Some concerns | Some concerns | Low | Low | Low | Some concerns |
| Phillips 2019 | Some concerns | Low | High | Low | Low | High |
| Polley 2002 | Some concerns | High | High | Low | Low | High |
| Poston 2015 | Some concerns | Low | High | Low | Low | High |
| Price 2012 | Low | Low | High | Low | Low | High |
| Quinlivan, 2011 | Low | Low | Low | Low | Some concerns | Some concerns |
| Renault, 2014 | Low | High | High | High | Low | High |

| | | | | | | |
|---|---|---|---|---|---|---|
| Ruiz, 2013 | Some concerns | Low | Low | Low | Low | Some concerns |
| Sagedal, 2017 | Low | Low | Low | Low | Low | Low |
| Sahariah， 2016 | Low | Low | High | Low | Low | High |
| Sales, 2018 | Some concerns | High | Low | Low | Low | High |
| Santamaria, 2015 | Some concerns | High | High | Low | Low | High |
| Seneviratne, 2015 | Low | Some concerns | Low | Low | Low | Some concerns |
| Simmons2017 | Low | Low | Low | Low | Low | Low |
| Stafne 2012 | Low | Low | Low | Low | Low | Low |
| Sun 2020 | Low | Low | Low | Some concerns | High | High |
| Syngelaki, 2016 | Low | Low | Low | Low | Low | Low |
| Thoronton 2009 | Low | Low | Low | Low | Some concerns | Some concerns |
| Trak-Fellermeier, 2019 | Low | Low | Low | Low | High | Some concerns |
| Valdés 2018 | Low | High | Low | Low | High | High |
| Van Horn, 2018 | Low | Low | Low | Low | Some concerns | Some concerns |
| Vanky 2010 | Low | Low | Low | Low | Some concerns | Some concerns |
| Vesco, 2014 | Low | Some concerns | Low | Low | Low | Low |
| Vinter, 2011 & 2014 | Low | Low | Low | Low | Low | Low |
| Vitale, 2021 | Low | High | Low | Low | Low | High |
| Walsh, 2012 | Low | Some concerns | Low | Low | Low | Some concerns |
| Wang, 2019 | Low | Some concerns | Low | Low | Low | Some concerns |
| Wickens 2017 | Low | Low | Low | Low | Low | Low |
| Wolff, 2008 | Low | Low | Low | Low | Low | Low |
| Xu, 2022 | Low | High | Low | Low | Low | Some concerns |
| Zhang, 2019 | High | High | High | Some concerns | Some concerns | High |
| Zhang, 2019 | Low | Low | Low | Low | Low | Low |
| Zhao, 2022 | Low | Low | Low | Low | Low | Low |
| D'Anna 213 | Some concerns | Low | Low | Low | Some concerns | Some concerns |
| Liu, 2021 | Low | Some concerns | Low | High | Some concerns | High |

Supplementary Table 5b: Risk of bias of cluster randomised controlled trials

| Author, year | Randomization process | Risk of bias arising from the timing of identification or recruitment of participants | Deviations from intended interventions | Missing outcome data | Measurement of the outcome | Selection of the reported result | Overall bias |
|---|---|---|---|---|---|---|---|
| Garmendia, 2020 | High | Low | Some concerns | Low | Low | Low | High |
| Hajian, 2020 | Some concerns | Low | Low | Low | Low | Some concerns | Some concerns |
| Kunath, 2019 | Low | Low | Low | Low | Some concerns | Some concerns | Some concerns |
| Lovvik, 2019 | Low | Low | High | Low | Low | Some concerns | Some concerns |
| Luoto, 2011 | Low | High risk | Low risk | Low | Low risk | Low risk | High |
| Rauh 2013 | High | Low | Low | Low | Low | Low | High |
| Wang, 2015 | Low | Some concerns | Low | Low | Low | Low | Some concerns |

Supplementary Table 5c: Risk of bias of non randomised controlled trials

| Author year | Bias due to confounding | Bias in selection of participants into the study | Bias in classification of interventions | Bias due to deviations from intended interventions | Bias due to missing data | Bias in measurement of outcomes | Bias in selection of the reported result | Overall bias |
|---|---|---|---|---|---|---|---|---|
| Opie 2016 | Critical | Low | Low | Critical | Low | Moderate | Low | Critical |
| Abd El Hameed, 2011 | Low | Low | Low | Moderate | Serious | Moderate | Low | Moderate |
| Ainuddin, 2015 | Serious | Moderate | Low | NI | Low | Low | Moderate | Serious |
| Epel 2019 | Critical | Low | Low | Critical | Low | Low | Low | Critical |
| Glueck 2002 | Critical | Low | Low | Critical | Critical | Moderate | Low | Critical |
| Glueck, 2002 | Serious | Low | Low | NI | Low | Low | Moderate | Serious |
| Gray-Donald, 2000 | Serious | Low | Low | NI | Low | Low | Low | Serious |
| Gregory, 2016 | Moderate | Low | Moderate | NI | Low | Low | Low | Moderate |
| Huvinen 2018 | Low | Low | Low | Moderate | Moderate | Moderate | Low | Moderate |
| Khattab, 2011 | Serious | Low | Serious | NI | NI | Low | Moderate | Serious |
| Liu, 2021 | Moderate | Low | Low | Moderate | Low | Moderate | Low | Serious |
| Shirazian, 2010 | Low | Low | Low | Low | Moderate | Low | Low | Moderate |
| Shirazian, 2014 | Low | Low | Low | Serious | Low | Low | Low | Serious |
| Sun, 2016 | Low | Low | Low | NI | Low | Low | Low | Low |
| Tomic 2013 | Critical | Low | Moderate | Low | Moderate | Moderate | Low | Critical |

Supplementary Table 6: GRADE summary

| **Intervention type** | **Risk of bias** | **Inconsistency** | **Indirectness** | **Imprecision** | **Publication bias** | **Certainty** | **Downgrade explanations** |
|---|---|---|---|---|---|---|---|
| Diet | Serious | Not serious | Not serious | Not serious | None | Moderate | Most studies have Some Concerns or High risk of bias |
| Physical activity | Serious | Not serious | Not serious | Not serious | None | Moderate | Most studies have Some Concerns or High risk of bias |
| Diet and physical activity | Serious | Serious | Not serious | Not serious | None | Low | Most studies have Some Concerns or High risk of bias;<br>High levels of heterogeneity |
| Metformin | Serious | Very serious | Not serious | Not serious | Present | Very low | Most studies have Some Concerns or High risk of bias;<br>High levels of heterogeneity;<br>Egger test suggests significant publication bias |
| Myoinositol | Serious | Very serious | Not serious | Not serious | None | Very low | Most studies have Some Concerns or High risk of bias;<br>Very high levels of heterogeneity |
| Probiotics | Serious | Very serious | Not serious | Serious | None | Very low | Most studies have Some Concerns or High risk of bias;<br>Very high levels of heterogeneity;<br>Pooled CI crosses 1.0 suggesting imprecision |

Supplementary Figure 1. Funnel plots for lifestyle, metformin and dietary supplement interventions on GWG

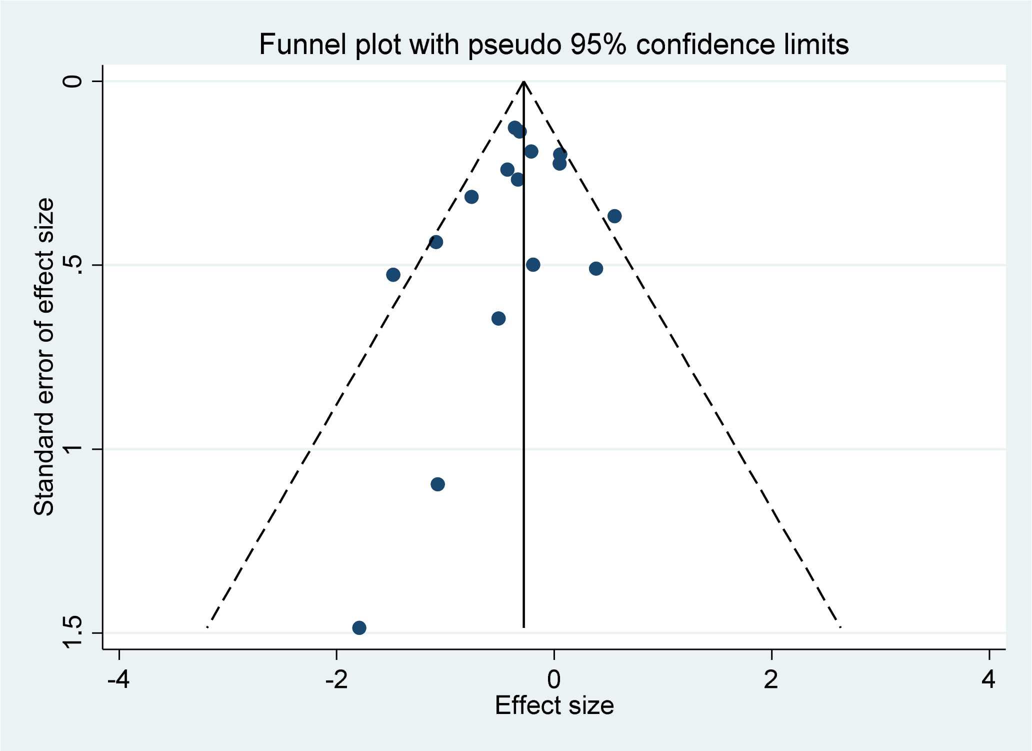


a) Diet

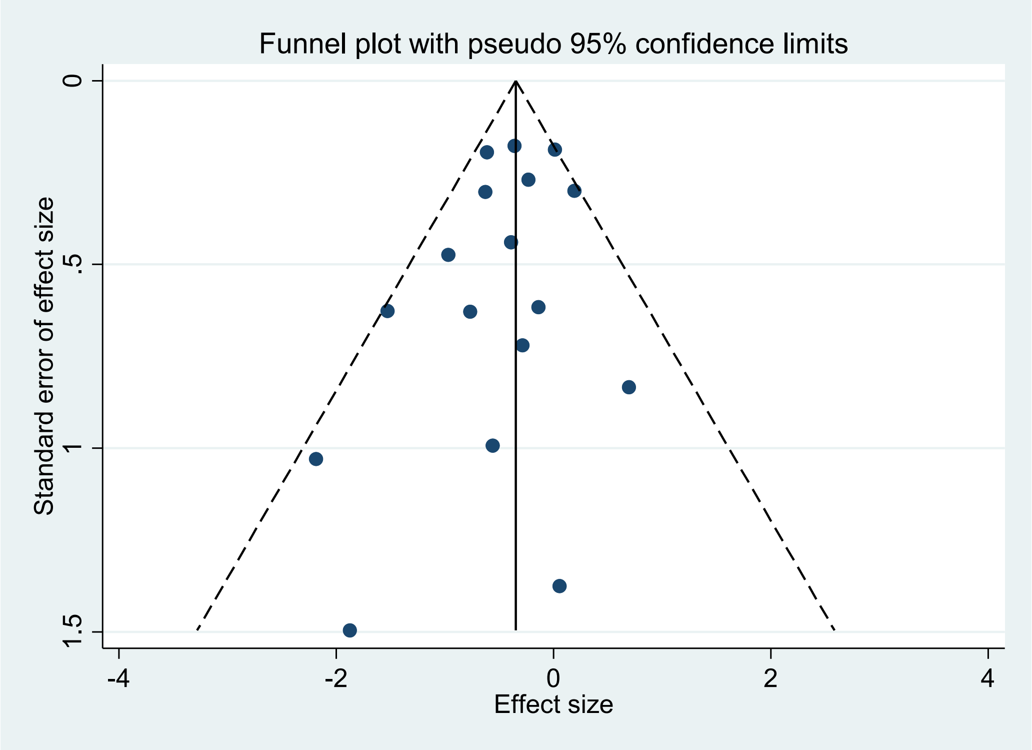


b) Physical activity

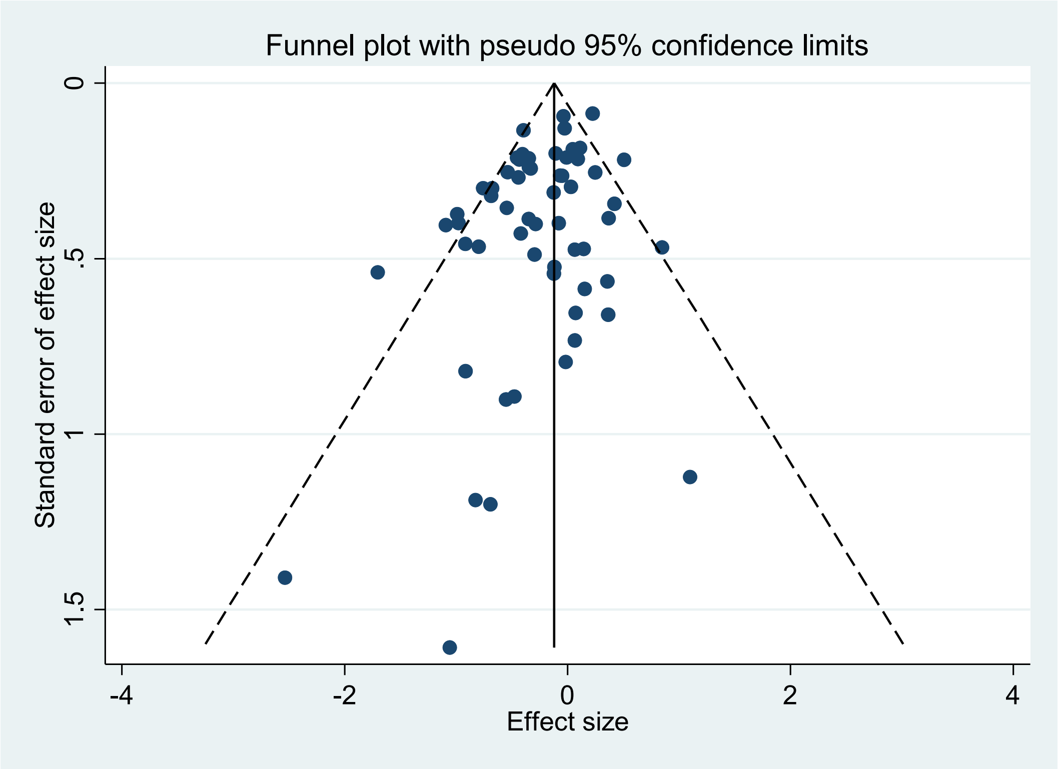


c) Combined diet and physical activity

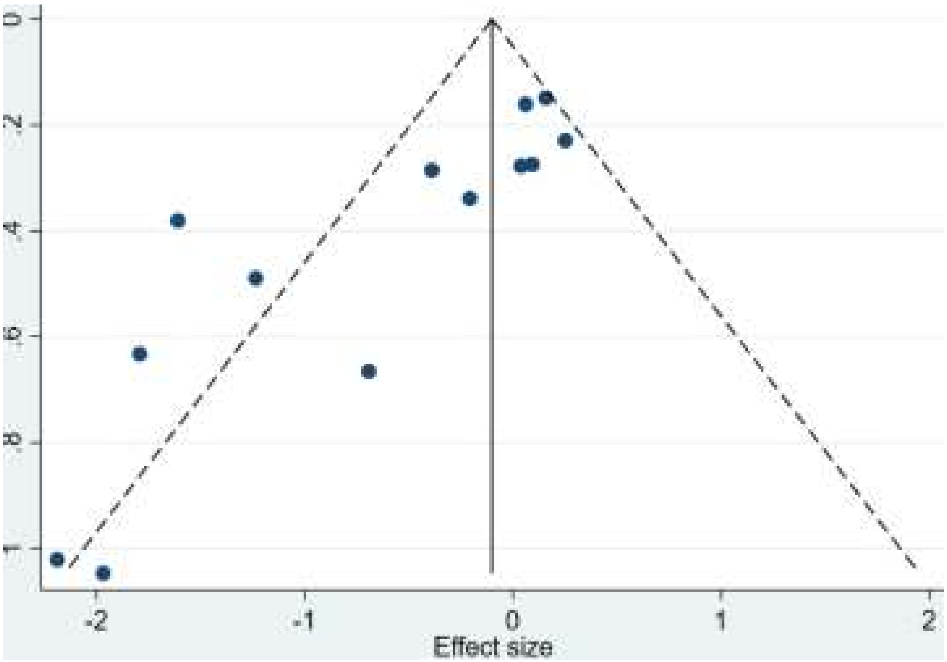


d) Metformin

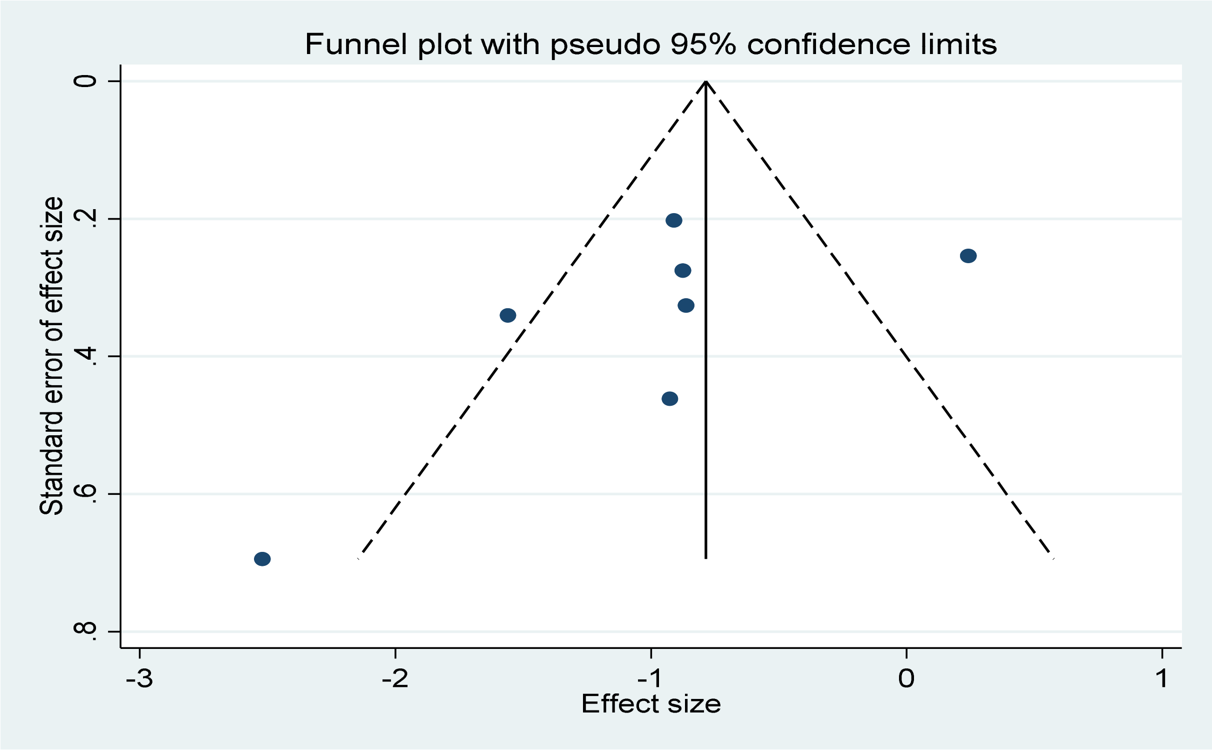


e) Myoinositol/inositol

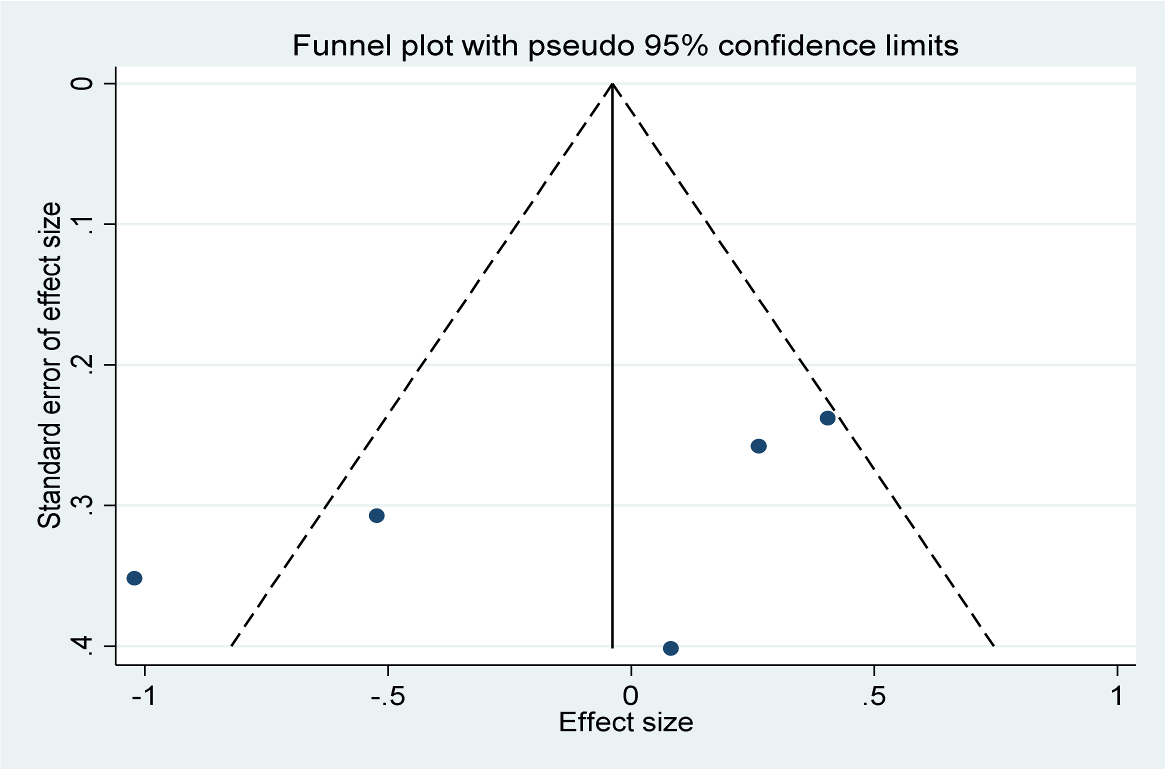


f) Probiotics
